# Geographic variation in the treatment of spinal disorders: association with health care professional availability, and population socioeconomic status, race, and ethnicity. A retrospective cohort study

**DOI:** 10.1101/2022.08.15.22278722

**Authors:** David Elton, Meng Zhang, Amy Okaya

## Abstract

**Importance:** Spinal disorders are common and associated with high rates of low value care. Efforts to improve guideline concordance and value benefit from understanding the influence of population and environmental factors.

**Objective:** Quantify geographic variation in population race/ethnicity, socioeconomic status, and health care professional (HCP) availability, and associated variation in service utilization and total cost for management of spinal disorders.

**Design, Setting, and Participants:** This retrospective cohort study examines a national sample of 1,534,280 complete episodes of a spinal disorder experienced in 2017-2019 using zip code and episode of care. Risk ratio (RR) and 95% confidence interval (CI), and ridge regression were used to examine associations between independent and dependent measures.

**Exposures:** Zip code level measures of population race/ethnicity and socioeconomic status were the primary independent measures.

**Main Outcomes and Measures:** Availability of and access to 17 types of HCPs, use of 14 types of health care services, and total cost were the primary dependent measures.

**Results:** 1,075,204 continuously insured individuals aged 18 years and older from 29,318 zip codes were associated with 1,534,280 episodes of a spinal disorder involving 531,115 HCPs generating $2,022,124,695 in expenditures. Compared to those in primarily white, middle income zip codes, individuals in non-white, disadvantaged zip codes were more likely to initially contact an emergency medicine physician (RR 2.23, 95% CI 2.11-2.36) or primary care provider (RR 1.40, 95% CI 1.38-1.42) and less likely to contact a chiropractor (RR 0.36, 95% CI 0.34-0.37). These individuals had higher exposure to prescription NSAIDs (RR 1.99, 95% CI 1.95-2.02), skeletal muscle relaxants (RR 1.57, 95% CI 1.52-1.59), opioids (RR 1.18, 95% CI 1.15-1.20), and CT scans (RR 1.94, 95% CI 1.84-2.04).

**Conclusions and Relevance:** Individuals in affluent, primarily non-Hispanic white zip codes have an abundance of options for managing a spinal disorder. Use of first line non-pharmacological and non-interventional options should be reinforced before second- and third-line services are considered. Individuals in low-income, non-white zip codes have less availability of non-pharmacological options, leading to greater use of emergency department and primary care and resulting in pharmaceutical management of spinal disorders. Sustainable models that increase availability of non-pharmaceutical options warrant further study.

**Key Points:** *Question:* Compared to the middle-income zip codes with a primarily non-Hispanic white population, do individuals with a spinal disorder living in zip codes characterized by socioeconomic disadvantage and a primarily non-white population have different care experiences?

*Findings:* Individuals with low back pain living in non-white, low-income zip codes have less availability of chiropractors, physical therapists and licensed acupuncturists and are more likely to seek initial treatment from an emergency department or primary care provider. These same individuals are more likely to receive pharmaceutical management, including prescription opioids, and less likely to receive guideline recommended non-pharmaceutical and non-interventional first line treatments.

*Meaning:* These findings suggest the need to create economically sustainable models that ensure access to guideline concordant non-pharmaceutical treatment options in non-white, low-income communities.

## Background

Spinal disorders are prevalent^1^, costly^2^, and a common reason for visiting a primary care provider (PCP).^3-5^. LBP clinical practice guidelines (CPGs)^6,7^ describe a stepped approach to management^3,8,9^ in which favorable natural history, self-care and non-pharmaceutical services are recommended as first-line approaches. Non-guideline concordant management of LBP is associated with a large percent of low-value care^3,10-14^ and with the risk of LBP transitioning from an acute to chronic condition.^15^ The likelihood of receiving CPG concordant management of LBP and neck pain (NP) has been associated with the type of initial health care professional (HCP) contacted.^3,15-22^

Geographic variation in health care quality, cost and outcomes is well established^23-32^ for a range of conditions, which is determined in some measure by both supply and demand factors.^33,34^ Socioeconomic status (SES),^35^ health care professional availability,^34^ and population demographics^27,36^ contribute to disparities in healthcare delivery and outcomes. A variety of geographic units of measurement are available to explore geographic variation,^37-42^ with tradeoffs identified with each approach.^43,44^

Geographic variation in the management of LBP has been described. The availability of different types of HCPs managing LBP has been shown to have an impact on cost^45,46^ and on use of health care services^47-49^. A variety of factors are associated with variation in the availability of, access to and utilization of first line non-pharmaceutical and non-interventional services^50,51^.

The aim of this study was to contribute to the understanding of geographic variation in the management of spinal disorders through analysis of a national sample of commercially insured adults in the US. The hypothesis was that HCP availability, service utilization, and total cost would vary based on population SES, race, and ethnicity.

## Methods

### Study design, population, setting and data sources

This is a retrospective cohort study of individuals seen by one or more HCPs for a complete episode of a spinal disorder. An analytic database was created by linking multiple databases. An enrollee database included de-identified individual data, and administrative claims data for all inpatient, outpatient, and pharmacy services. An HCP database consisted of de-identified HCP demographic information and professional licensure status.

Because data was linked from various sources, a review was performed to assess compliance with de-identification requirements. With data being de-identified or a Limited Data Set in compliance with the Health Insurance Portability and Accountability Act, the UnitedHealth Group Office of Human Research Affairs determined that Institutional Review Board approval or waiver of authorization was not required. The study was conducted and reported based on the Strengthening the Reporting of Observational Studies in Epidemiology (STROBE) guidelines (Supplement – STROBE Checklist).^52^

### Unit of analysis and cohort selection

Episode of care and 5-digit zip code were the units of analysis. Episodes have been shown to be a valid way to organize administrative claims data.^53^ The *Symmetry*^*®*^ *Episode Treatment Groups*^*®*^ *(ETG*^*®*^) and *Episode Risk Groups*^*®*^ *(ERG*^*®*^) version 9.5 methodologies and definitions were used to translate administrative claims data into discrete episodes.^54^

Complete episodes were defined as having at least 91-day pre- and 61-day post-episode clean periods during which no services were provided by any HCP for any LBP or NP diagnosis. Incomplete episodes were excluded. All non-musculoskeletal episodes and musculoskeletal episodes not categorized as LBP or NP were excluded. LBP and NP episodes associated with diagnoses of malignant and non-malignant neoplasms, fractures and other spinal trauma, infection, congenital deformities and scoliosis, autoimmune disorders, osteoporosis, and advanced arthritis were excluded.

The home address 5-digit zip code of the individual with LBP or NP was used as the crosswalk between study data sources. HCP location was based on the 5-digit zip code of the office location where a service was provided. HCP zip codes associated with a location without population data were transformed to the geographically closest zip code with material population. Population size, race, and ethnicity was obtained from the US Census Bureau^55^ using the zip code tabulation area (ZCTA) code. Household adjusted gross income (AGI)^56^ was obtained from the Internal Revenue Service using the 5-digit zip code of the filer home address. Area Deprivation Index (ADI)^57^ was obtained from the University of Wisconsin Neighborhood Atlas at the zip+4 level and was transformed to a 5-digit zip code.

The cohort consisted of individuals aged 18 years and older with a complete episode of LBP or NP commencing and ending during the calendar years 2017-2019. This timeframe was selected as the 3-year period before the influence of the COVID-19 epidemic in early 2020. All individuals had continuous medical and pharmacy coverage from a single national insurer during the entire study period.

### Variables

Python (*Python Language Reference, Version 3*.*7*.*5*., n.d.) was used for data preprocessing, table generation, and initial analyses. R (*version 3*.*6*.*1*) was used for Ridge regression analyses. A goodness-of-fit analysis was conducted using D’Agostino’s K-squared test. Non-normally distributed data were reported using median and interquartile range (IQR).

The primary independent variables were zip code ADI and population percent non-Hispanic white (NHW) with each variable segmented into 4 categories, and 16 combined ADI/NHW segments. The reference for analyses of statistical significance was the highest episode volume 25-50 ADI and 75-100% NHW segment.

### Measures

The number of HCPs per 1,000 population was calculated for 17 types of HCP commonly contacted initially for LBP or NP. Doctors of Osteopathy (DO) with evidence of utilizing manipulative treatment were separately reported, and otherwise were included in the HCP type for which the DO was boarded. A Primary Care Provider (PCP) category consisted of Family Practice, Internal Medicine, General Medicine and OBGYN physician types. The Nurse category consisted primarily of nurse practitioners. Medical physician types not separately reported were reported as “MD-other”.

The count of unique HCPs within a 5-digit zip code was based on all in- and out-of-network HCPs who provided at least one service of any type for any individual with a neuromusculoskeletal (NMS) condition during the 2017-2019 study period. The larger NMS cohort included a wide range of conditions in addition to LBP and NP. The HCP zip code was based on the office location where a service was provided. Basing the HCP count on the larger NMS sample helped minimize the potential for misclassification bias associated with under-counting available HCPs.

The percent of episodes where an individual initially contacted each HCP type was calculated. All HCP types could be accessed directly without a referral and the HCP office location could be within or outside the individual’s home address zip code. The rate and timing of use of 14 types of health care services was calculated.^3,4^ Service utilization reflects services provided at any time by any type of HCP. Services were grouped into first-, second-, and third-line categories. First-line services include chiropractic and osteopathic manipulation, active care, manual therapy, and acupuncture. Second-line services include prescription NSAIDs and skeletal muscle relaxants, radiography, and MRI studies. Third-line services include prescription opioids, spinal injection and surgery, and CT scans. Odds (OR) and risk (RR) ratios, along with associated 95% confidence intervals (CI), were calculated for both the initial contact HCP and utilization of each service type. As a measure more widely understood in associational analyses, RR were reported.^58^

The total cost, number of different HCPs seen during an episode, and episode duration were calculated. Total cost included costs associated with all services provided by any HCP for a LBP or NP episode, including those not specifically identified in the 14 categories used in the analyses. Costs for services for which an insurance claim was not submitted were not available. Episode duration was the number of days between the first and last date of service for each episode.

Due to potential collinearity among HCPs per 1000 and initial contact HCP measures, ridge regression was used to better identify significant predictors among zip code population factors, HCP availability, and dependent measures. Variables were scaled to create similar ranges. ADI was transformed from a 0-100 scale to 0-1, and HCP type per 1000 was transformed to per 100. Log transformed, and non-transformed total cost results were produced in addition to opioid and NSAID prescribing rates using a 0-1 scale.

## Results

The sample included 1,075,204 individuals, with a median age of 44.0 (Q1 34.0, Q3 54.0) and 54.9% females. These individuals were from all 50 States, 29,318 5-digit zip codes, and were associated with 1,534,280 complete episodes of a spinal disorder involving 531,115 unique HCPs and $2,022,124,695 in reimbursed health care expenditures. The median pre-episode clean period was 621 days (Q1 409, Q3 853). For individuals with multiple episodes the median number of days between sequential episodes was 202 (Q1 120, Q3 336). The median post-episode clean period was 435 days (Q1 266, Q3 690). (Supplement 1) Among the ADI and percent NHW segments, the distribution of LBP and NP episodes, individual age, % female, ERG® risk score, and number of different HCP seen was highly uniform. (Supplement 2)

### Availability of HCPs

Primary care HCP types were available across ADI segments median 1.06 per 1000 population (Q1 0.42, Q3 2.38), with the lowest availability observed in zip codes with 0-25% NHW population (median 0.59, Q1 0.25, Q3 1.34). Availability of non-prescribing (median 0.45, Q1 0.18, Q3 0.88), specialist (median 0.25, Q1 0.03, Q3 1.01) and emergency/radiology/urgent care (EM/UC) (median 0.10, Q1 0.00, Q3 0.31) HCP types was lower than primary care, with availability of non-prescribing and specialist types increasing as ADI decreased. Like primary care, availability of non-prescribing (median 0.14, Q1 0.04, Q3 0.30) and specialist (median 0.11, Q1 0.01, Q3 0.49) HCP types were generally lowest in zip codes with 0-25% NHW population. (Supplements 3 and 3a)

### Initial Contact HCP

Zip codes with 75-100 ADI and 0-25% NHW population were associated with a higher percent of individuals initially contacting a primary care HCP (59.3%), and lower percent of individuals initially contacting a non-prescriber (16.9%) or specialist HCP (13.1%). As ADI decreased and NHW% increased the percent of individuals initially contacting a non-prescriber or specialist increased. [Figure 1] (Supplement 4)

**Figure 1.**
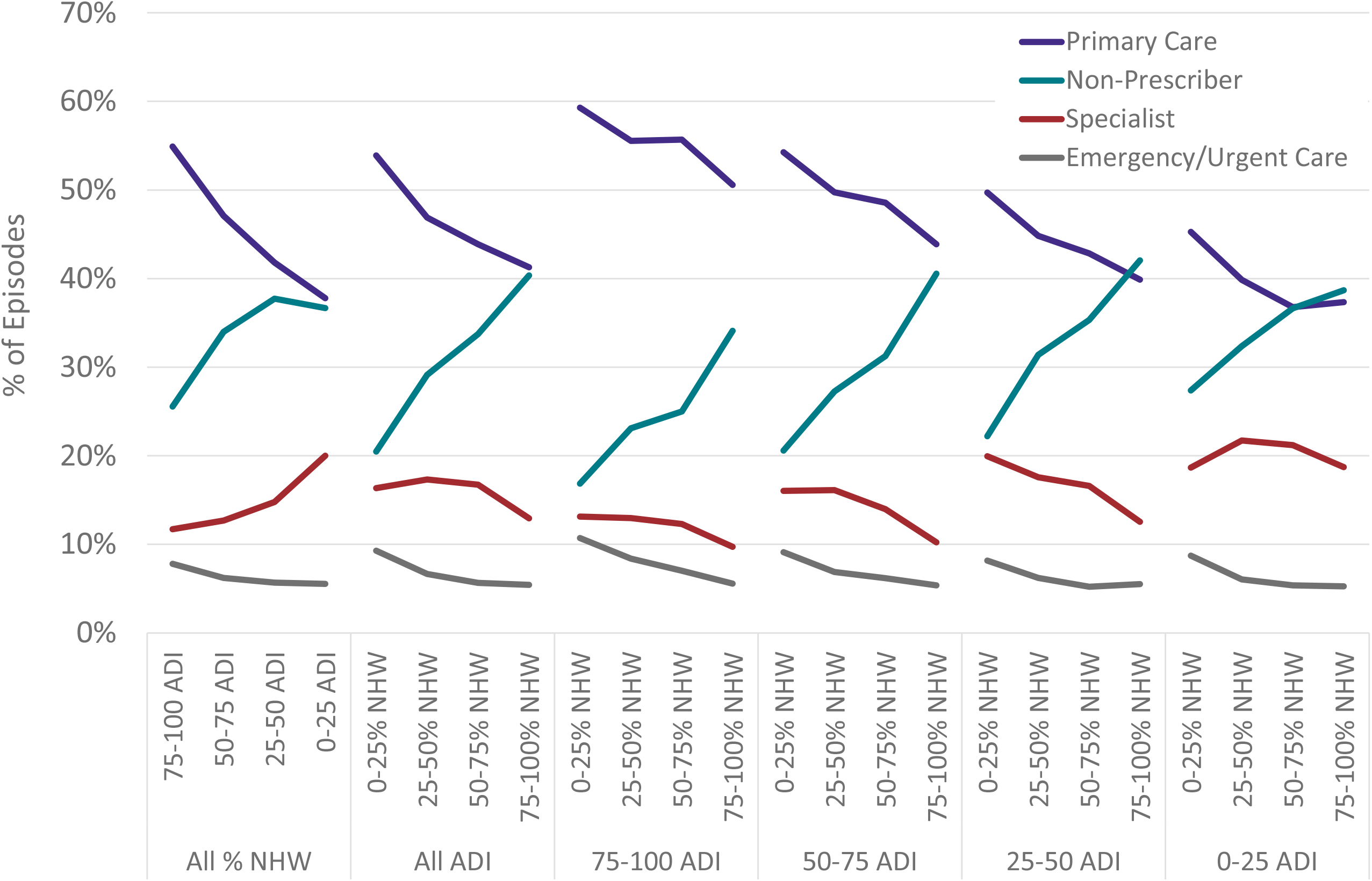
% of spinal disorder episodes with initial contact with primary care, non-prescriber, specialist and emergency medicine/urgent care health care providers segmented by individual home address zip code Area Deprivation Index (ADI) and percent non-Hispanic (NHW) white population

Compared to the reference segment, individuals with a spinal disorder in the 75-100 ADI and 0-25% NHW segment were more likely to contact an EM/UC HCP (RR 1.73, 95% CI 1.66-1.80), or primary care HCP (RR 1.34, 95% CI 1.32-1.35), and less likely to contact a non-prescribing HCP (RR 0.36, 95% CI 0.35-0.37). Individuals with a spinal disorder in the 0-25 ADI and 75-100% NHW segment were more likely to contact a specialist HCP (RR 1.42, 95% CI 1.39-1.44). [Figure 2] (Supplement 4a)

**Figure 2.**
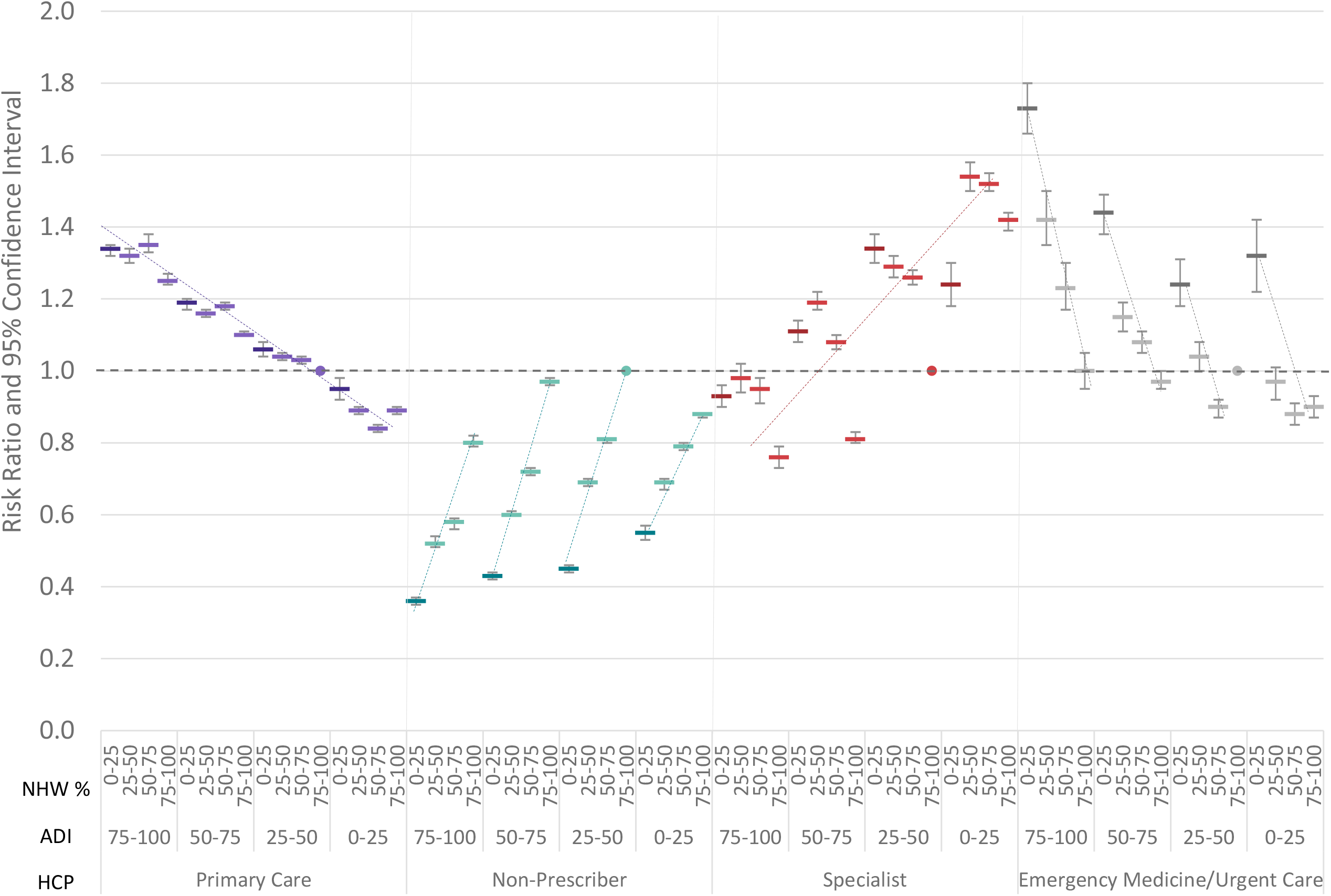
Risk ratio and 95% confidence interval for spinal disorder type of initial contact health care provider by 5 digit zip code population Area Deprivation Index (ADI) and % non-Hispanic white (NHW%) segementation compared to reference segment of 25-50 ADI and 75-100% NHW

### Services Provided for Spinal Disorders

As ADI decreased and the NHW% increased, second-line services decreased, and first-line services increased. Third-line service use had a stronger association with ADI than NHW%, decreasing as ADI decreased. The 75-100 ADI and 0-25% NHW segment was associated with second-line services (66.6% of episodes) and third-line services (24.0%) provided more frequently than first-line services (23.4%). The 0-25 ADI and 75-100% NHW segment was associated with first-line services provided more frequently (52.2%) than second-line services (47.8%), or third-line services (19.0%). [Figure 3] (Supplement 5)

**Figure 3.**
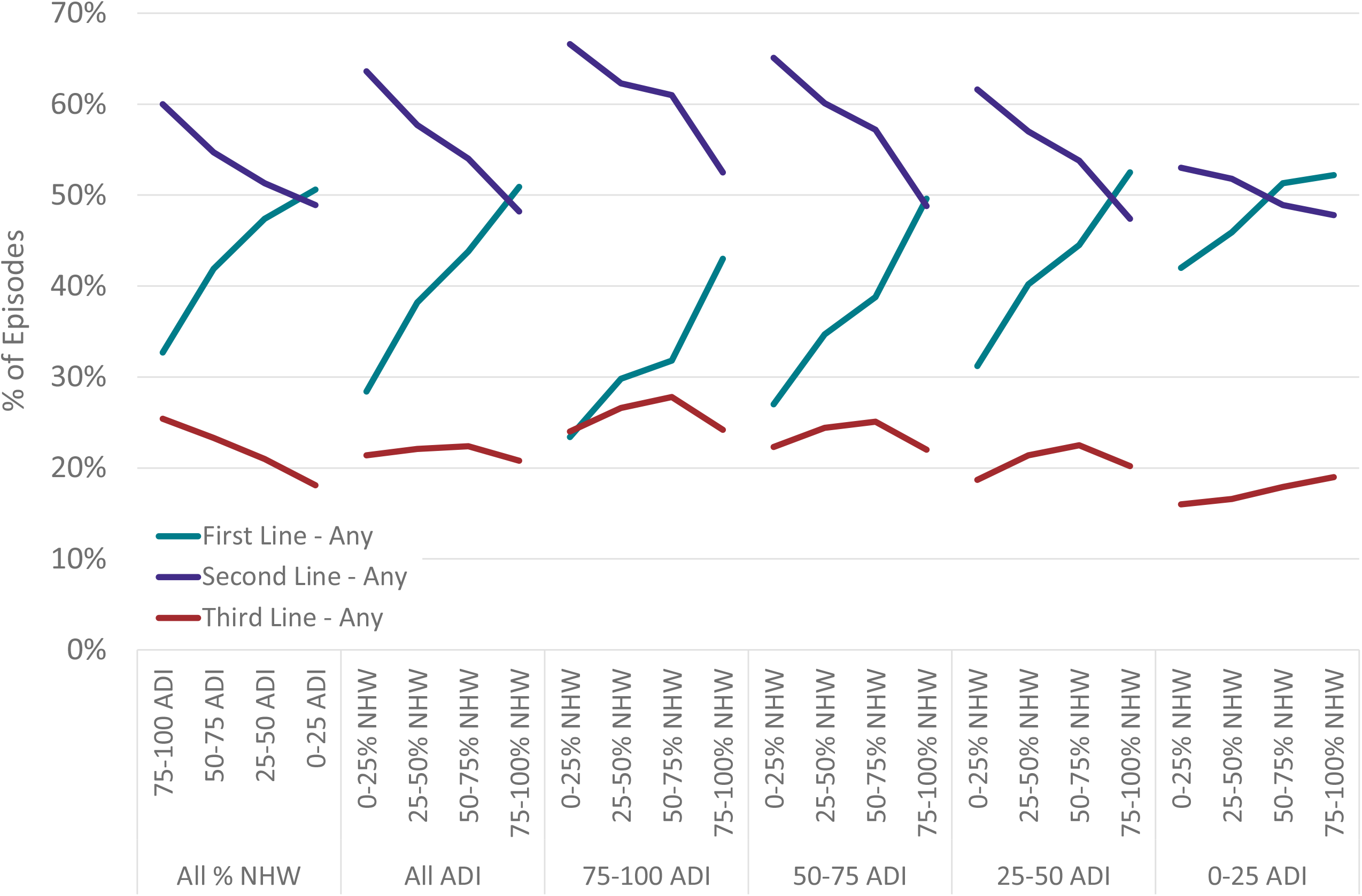
% of spinal disorder episodes including first, second and third line services segmented by individual home address zip code Area Deprivation Index (ADI) and percent non-Hispanic (NHW) white population

Compared to the reference segment, individuals in the 75-100 ADI and 0-25% NHW segment were more likely to receive a second-line service (RR 1.41, 95% CI 1.40-1.42) or a third-line service (RR 1.19, 95% CI 1.16-1.21), and less likely to receive a first-line service (RR 0.45, 95% CI 0.44-0.45). [Figure 4] Compared to the reference segment, individuals in the 75-100 ADI and 0-25% NHW segment were more likely to receive a prescription NSAID (RR 1.99, 95% CI 1.95-2.02) and less likely to receive chiropractic manipulation therapy (CMT) (RR 0.34, 95% CI 0.33-0.35) or active care (RR 0.68, 95% CI 0.66-0.70). Compared to the reference segment, individuals in the 75-100 ADI and 50-75% NHW segment were more likely to receive a prescription opioid (RR 1.47, 95% CI 1.44-1.51). Within each ADI segment CMT and prescription NSAIDs had a strong association with NHW%. [Figure 5] (Supplement 5a) There were statistically significant, yet not clinically meaningful differences in the timing of introduction of most health care services between ADI and NHW segments. (Supplement 6)

**Figure 4.**
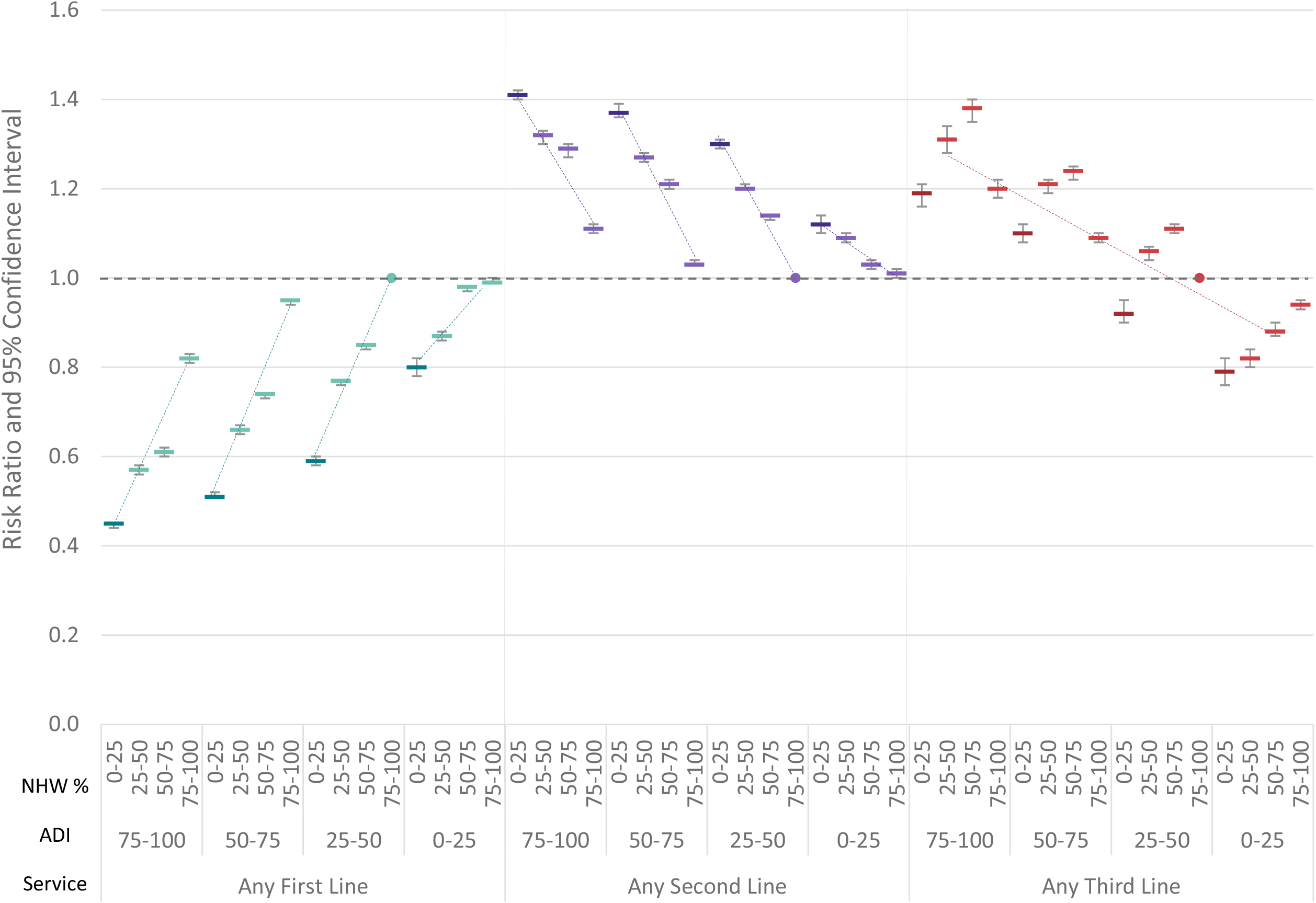
Risk ratio and 95% confidence interval for spinal disorder service use by 5 digit zip code population Area Deprivation Index (ADI) and % non-Hispanic white (NHW%) segementation compared to reference segment of 25-50 ADI and 75-100% NHW

**Figure 5.**
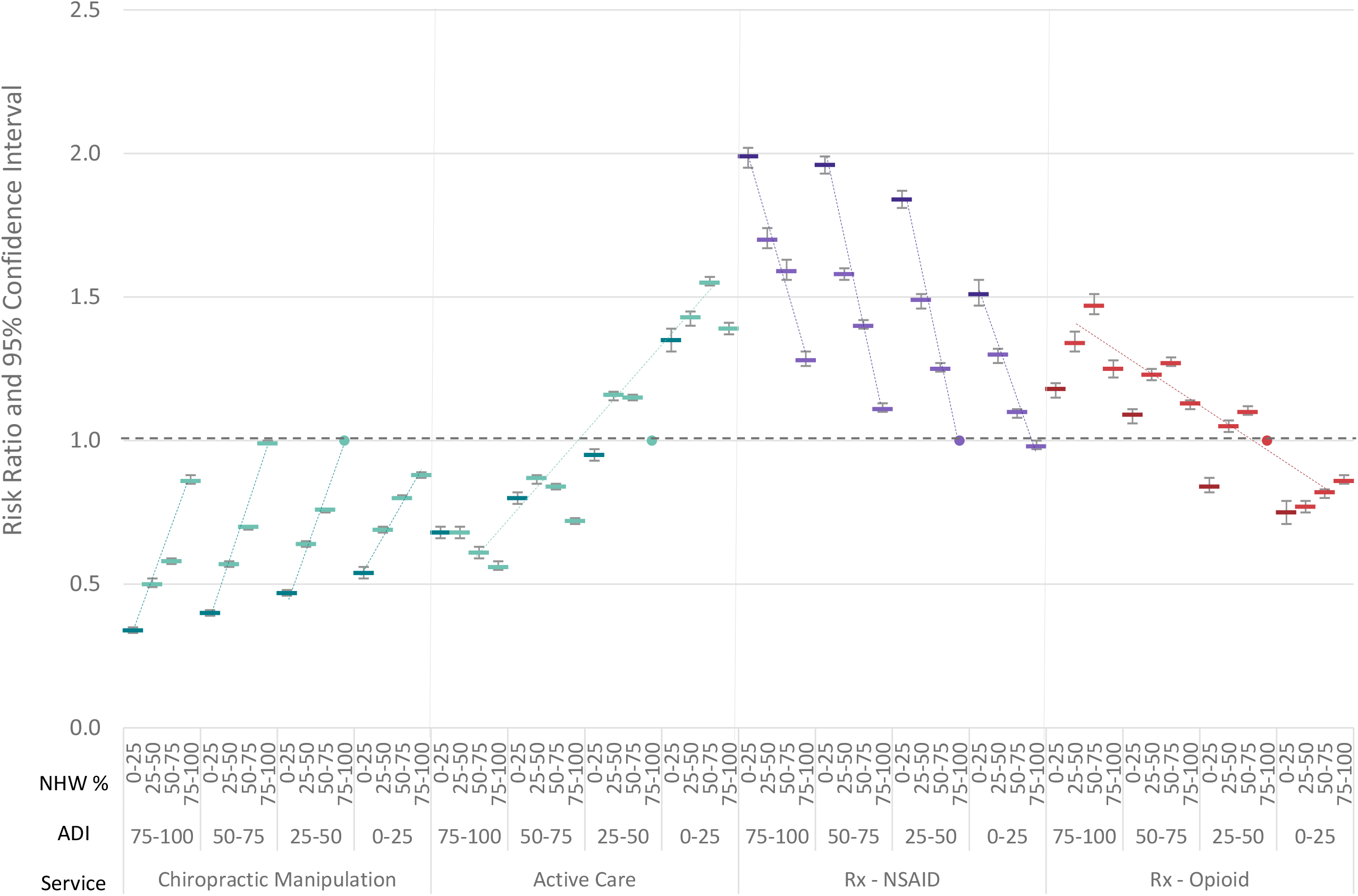
Risk ratio and 95% confidence interval for spinal disorder service use by 5 digit zip code population Area Deprivation Index (ADI) and % non-Hispanic white (NHW%) segementation compared to reference segment of 25-50 ADI and 75-100% NHW

### Episode Cost

Total cost increased as ADI decreased. (Supplement 7) Episodes in zip codes with high ADI and low NHW% had lower total cost associated with higher rates of pharmacological services, lower rates of first line non-pharmacological services, and lower rates of spinal surgery. Zip codes with low ADI and high NHW% population had higher total cost associated with lower rates of pharmacological services, higher rates of first-line non-pharmacological services, higher rates of MRI and spinal injection.

Ridge regression revealed many favorable (-) and unfavorable (+) associations with selected dependent measures. Notably, a higher number of DCs per 100 population was associated with a lower rate of opioid exposure, while a higher number of Pain Management physicians was associated with a higher rate of opioid exposure. The type of HCP initially contacted by an individual with a spinal disorder was associated with a significant difference in the rate of opioid exposure. Zip codes with higher ADI were associated with a higher rate of opioid prescribing while zip codes with a higher % non-Hispanic Asian population were associated with lower rates of opioid exposure. (Supplements 8 and 8a)

## Discussion

This study provides a comprehensive analysis of associations between population SES and race/ethnicity, and the management of spinal disorders. Zip codes with higher ADI, and a primarily non-white population, were associated with less availability of all types of HCP, particularly those providing guideline concordant first-line treatment options. Individuals with a spinal disorder in these zip codes were more likely to initially contact PCPs, nurses, and emergency medicine physicians, which was associated with greater use of prescription medications, including opioids. Individuals in zip codes with low ADI, and a primarily NHW population, were associated with more abundant availability of, access to and utilization of a range of HCPs and services.

This study has several potential limitations. While administrative data benefited from extensive quality and actuarial control measures, variability in benefit plan design and enrollee cost-sharing responsibility, errors and missing information were potential sources of confounding or bias. Similarly, HCP type and office location data may have included errors or missing information. Lack of data about individual and HCP race/ethnicity prevented exploration of individual preferences for sex/racial/ethnic concordance when selecting an initial HCP. The data set did not describe a U.S representative sample.

5-digit zip code was selected as a reasonable approach to balancing limitations of each geographic unit of measurement. County- or state-level measures blur meaningful environmental and population factors. Census tract or census block groups, while improving precision of population factors, introduces limitations in evaluating HCP availability and raises potential individual confidentiality risks. Associating 5-digit zip code with the corresponding ZCTA is subject to minor risk of misclassification bias. Transforming ADI zip+4 to a 5-digit zip code is associated with potential misclassification bias. This was partially mitigated by converting ADI data to categorical data for use as an independent variable.

Calculating the number of HCPs per 1,000 population is subject to both selection and misclassification bias. The HCP count was based on the number of unique in or out of network HCPs providing at least one service for any NMS condition during the three-year study period from an office location within the same zip code as the individual with a spinal disorder. Missing from this count are all possible HCPs with an office location in the zip code of interest.

There are potential limitations associated with using episode of care as the unit of analysis. As demonstrated in earlier studies of the same underlying data the risk of episode misclassification bias is low.^3,4^ Further minimizing this risk was the assumption that any misclassification bias associated with episode type would be similar within each ADI and percent NHW segment.

A potential translation limitation of this study is that the results were derived from a commercially insured population with continuous coverage for a broad range of services available through direct access to a range of HCPs. Even in this best-case scenario, the management of spinal disorders was associated with meaningful disparities in availability of, access to and utilization of health care services. It is unclear how these observed patterns are amplified or muted in a population with less or no coverage for guideline-recommended first-line services.

This study expands on earlier studies of the same LBP and NP cohorts^3,4^ and contributes to the understanding of geographic variation in health care, and specifically variation in management of spinal disorders associated with environmental and population factors such as SES, race/ethnicity, and HCP availability.

Within each ADI segment, zip codes with 75-100 percent non-white population were associated with a 150-200% greater likelihood of prescription NSAID use for LBP or NP, the largest disparity identified in the study. With the potential adverse events associated with chronic NSAID use,^59^ this finding warrants further study, particularly with other studies not finding similar disparities in NSAID use.^60-62^

The geographic distribution of rate of opioid prescribing corroborates similar analyses.^63,64^ (Supplement – Maps) Previous studies of variation in opioid prescribing have found a weak association with both availability of prescribers and prescribing practices, and demand from individuals with pain.^48^ This study found the sparse availability of non-prescribing HCPs in areas of high ADI as a potentially important factor to consider in addressing opioid over-prescribing.

The strong inverse association between the availability of DCs and the rate of opioid prescribing, and the geographic distribution of DCs (Supplement – Maps), corroborates findings from other studies.^65,66^ Episodes in zip codes with a high percent NHW population were more likely have a DC as the initial contact HCP, corroborating similar findings.^67^ Similarly, this study corroborates the finding that if a PT or LAc is the initial contact HCP the rate of opioid exposure is lower.^3,4,17,68^ A previous study found that as part of a home exercise program PTs recommended fewer exercises for Black individuals.^69^ An association between population race/ethnicity and percent of episodes including active care was not found in this study; however, individuals living in zip codes with low ADI were more likely to receive active care than individuals living in zip codes with high ADI.

Higher rates of plain film radiography use were found in zip codes with higher ADI and lower percent of NHW population, and higher rates MRI use were found in zip codes with lower ADI and higher percent NHW population. A similar association between type of imaging study and population SES has been demonstrated in Canada for non-NMS conditions.^70,71^ Study findings differ from a study of MRI for acute work-related injury^71^ that found states with lower median income were associated with higher rates of early MRI use. Previous findings of geographic variation in rates of spinal injection,^72^ and spinal surgery^73,74^ were also found in this study.

## Conclusions

For individuals with LBP or NP, the zip code one lives in has a strong association with the availability and likelihood of receiving guideline concordant high-value care. Individuals living in zip codes with socioeconomic disadvantage and a low percent NHW population have lower availability of non-prescribing HCPs. For LBP and NP, this is associated with more use of primary care and emergency department HCPs. Not surprisingly, these same zip codes are associated with higher rates of prescription NSAID and opioid use. Economic factors in these zip codes may present a barrier to a non-prescribing HCP maintaining a viable solo or small practice. Further exploration of this potential barrier warrants further study. Individuals living in affluent zip codes with a high percent NHW population have an abundance of HCP options. While more likely to receive early first-line guideline concordant care, these individuals are also more likely to receive low-value care resulting in higher total episode cost. Similar to other conditions efforts to improve guideline concordance and value in the management of spinal disorders must consider local environmental and population factors.

## Supporting information

Supplement - STROBE Checklist

Supplement 1 - Cohort

Supplement 2 - Cohort By Segment

Supplement 3 - HCP Count

Supplement 3a - HCP Count Figure

Supplement 4 - Initial Contact HCP

Supplement 4a - Initial Contact HCP Risk Ratio

Supplement 5 - Services

Supplement 5a - Services Risk Ratio

Supplement 6 - Services Timing

Supplement 7 - Cost

Supplement 8 - Ridge Regression

Supplement 8a - Ridge Regression Figure

Supplement - Maps

## Data Availability

All data produced in the present work are contained in the manuscript

## Declarations

### Ethics approval and consent to participate

Because data was linked from various sources, a review was performed to assess compliance with de-identification requirements. With data being de-identified or a Limited Data Set in compliance with the Health Insurance Portability and Accountability Act and customer requirements, the UnitedHealth Group Office of Human Research Affairs determined that this study was exempt from Institutional Review Board review.

### Consent for publication

Not applicable

### Availability of data and materials

The data are proprietary and are not available for public use but, under certain conditions, may be made available to editors and their approved auditors under a data-use agreement to confirm the findings of the current study.

### Authors’ contributions

Study conception and design; **DE, MZ**. Data acquisition; **DE, MZ**. Data analysis and interpretation; **DE, MZ**. Draft or substantially revise manuscript; **DE, MZ and AO**.

## Acknowledgements

Thomas Kosloff, DC and Scott Shimotsu, PhD MPH made important contributions to the study conception, design, and early draft review.

## List of Abbreviations

LBP: Low back pain
PCP: Primary care provider
CPG: Clinical practice guideline
HCP: Health care professional
SES: Socioeconomic status
US: United States
AGI: Adjusted gross income
ADI: Area Deprivation Index
HL: Health literacy
STROBE: Strengthening the Reporting of Observational Studies in Epidemiology
ETG^*®*^: Episode Treatment Group^*®*^
ERG^*®*^: Episode Risk Group^*®*^
FIPS: Federal Information Processing Standard
ZCTA: zip Code Tabulation Area
ACP: American College of Physicians
NHW: Non-Hispanic White
NHB: Non-Hispanic Black
NHA: Non-Hispanic Asian
DO: Doctor of Osteopathy
OBGYN: Obstetrics and Gynecology
NMS: Neuromusculoskeletal
SD: Standard deviation
OR: Odds ratio
RR: Risk ratio
CI: Confidence interval
DC: Doctor of Chiropractic
PT: Physical Therapist
EM: Emergency Medicine
LAc: Licensed Acupuncturist
PA: Physician’s Assistant
MRI: Magnetic resonance imaging
CT: computed tomography
NSAID: Non-steroidal anti-inflammatory drug
CMT: Chiropractic manipulative treatment
OMT: Osteopathic manipulative treatment

