## Supplement 1 - Cohort for "Geographic variation in the treatment of spinal disorders: association with health care professional availability, and population socioeconomic status, race, and ethnicity. A retrospective cohort study"

| Supplement 1 - Cohort | |
|---|---|
| Zip Codes | 29318 |
| Individuals | 1075204 |
| Complete Episodes | 1534280 |
| Health Care Providers (HCP) | 531115 |
| Total Episode Cost | 2022124695 |
| **Episodes - Median (IQR)(Q1, Q3)** | |
| Total Episode Cost | 221 (618) (87, 705) |
| **Episode Timing - Median (IQR)(Q1, Q3)(Minimum)** | |
| Episode Durations (days) | 26 (133) (1, 134) (1) |
| Clean Period Before Initial Episode (days) | 621 (444) (409, 853) (91) |
| Clean Period Between Sequential Episodes (days) | 202 (216) (120, 336) (61) |
| Clean Period After Final Episode (days) | 435 (424) (266, 690) (61) |
| **Individuals - % or Median (IQR)(Q1, Q3)** | |
| Female % | 54.9% |
| Age (years) | 44.0 (20.0) (34.0, 54.0) |
| ERG® Risk Score | 1.5 (0.9) (1.1, 2.0) |
| Episodes Per Individual | 1.0 (1.0) (1.0, 2.0) |
| **Zip Code Characteristics - Median (IQR)(Q1, Q3)** | |
| Episodes per zip code - mean (SD) | 12.0 (48.0) (3.0, 51.0) |
