## Supplement 2 - Cohort By Segment for "Geographic variation in the treatment of spinal disorders: association with health care professional availability, and population socioeconomic status, race, and ethnicity. A retrospective cohort study"

### Supplement 2 - Cohort and population attributes by zip code Area Deprivation Index (ADI) and population % non-Hispanic white (NHW) segments

| ZIP code Area Deprivation Index (ADI) | ZIP code % Non-Hispanic White (NHW) population | Episodes | % of Episodes | % Low back pain (LBP) episodes | % Neck pain (NP) episodes | Individuals - % female | Individual age - Median (Q1, Q3) | Individual Episode Risk Group® (ERG®) score - Median (Q1, Q3) | ZIP code household Adjusted Gross Income (AGI) - Median (Q1, Q3) | ZIP code Area Deprivation Index (ADI) - Median (Q1, Q3) |
|---|---|---|---|---|---|---|---|---|---|---|
| Total | | 1534280 | 100.0% | 65.6% | 34.4% | 56.0% | 45 (34, 54) | 1.5 (0.7, 3.0) | 65,144 (50,843, 91,603) | 44.7 (27.4, 61.7) |
| 75-100 | All | 133334 | 8.7% | 68.3% | 31.7% | 55.4% | 45 (35, 54) | 1.6 (0.7, 3.2) | 39,836 (33,617, 45,025) | 80.5 (77.5, 84.6) |
| 50-75 | All | 502531 | 32.8% | 66.6% | 33.4% | 55.9% | 45 (34, 54) | 1.5 (0.7, 3.1) | 51,883 (46,185, 58,772) | 61.4 (55.6, 67.5) |
| 25-50 | All | 559487 | 36.5% | 65.1% | 34.9% | 56.2% | 45 (34, 54) | 1.4 (0.7, 2.9) | 72,840 (62,612, 88,304) | 37.9 (31.7, 43.9) |
| 0-25 | All | 325481 | 21.2% | 64.0% | 36.0% | 56.0% | 45 (35, 54) | 1.4 (0.7, 2.9) | 118,680 (94,064, 163,283) | 16.0 (10.3, 20.7) |
| All | 0-25 | 125698 | 8.2% | 67.7% | 32.3% | 58.0% | 44 (34, 54) | 1.5 (0.7, 3.0) | 40,620 (33,196, 50,146) | 63.8 (42.3, 76.7) |
| All | 25-50 | 224009 | 14.6% | 65.9% | 34.1% | 56.8% | 44 (34, 54) | 1.5 (0.7, 3.0) | 56,445 (45,523, 76,002) | 48.9 (29.1, 64.5) |
| All | 50-75 | 462283 | 30.1% | 65.1% | 34.9% | 55.9% | 45 (34, 54) | 1.5 (0.7, 3.1) | 71,641 (54,239, 103,665) | 41.0 (24.6, 57.2) |
| All | 75-100 | 702250 | 45.8% | 65.5% | 34.5% | 55.4% | 46 (35, 55) | 1.4 (0.7, 2.9) | 69,294 (55,356, 94,206) | 42.7 (27.1, 59.2) |
| 75-100 | 0-25 | 35387 | 2.3% | 69.1% | 30.9% | 57.6% | 45 (35, 54) | 1.5 (0.7, 3.1) | 31,438 (28,692, 34,159) | 82.5 (78.8, 86.3) |
| 75-100 | 25-50 | 24343 | 1.6% | 68.6% | 31.4% | 55.6% | 45 (35, 54) | 1.6 (0.7, 3.2) | 37,976 (34,676, 42,242) | 80.7 (77.4, 83.9) |
| 75-100 | 50-75 | 27185 | 1.8% | 68.0% | 32.0% | 54.5% | 46 (35, 55) | 1.7 (0.8, 3.3) | 42,155 (38,215, 46,454) | 79.4 (77.4, 83.5) |
| 75-100 | 75-100 | 41486 | 2.7% | 67.7% | 32.3% | 54.4% | 46 (36, 55) | 1.6 (0.7, 3.2) | 44,409 (40,715, 48,385) | 79.1 (76.9, 82.3) |
| 50-75 | 0-25 | 49075 | 3.2% | 68.2% | 31.8% | 58.3% | 44 (34, 54) | 1.5 (0.7, 3.0) | 39,953 (35,382, 46,119) | 64.9 (58.3, 70.6) |
| 50-75 | 25-50 | 84091 | 5.5% | 66.5% | 33.5% | 57.1% | 45 (34, 54) | 1.5 (0.7, 3.1) | 47,625 (42,656, 54,508) | 62.6 (56.7, 67.6) |
| 50-75 | 50-75 | 139453 | 9.1% | 66.3% | 33.7% | 55.7% | 45 (35, 54) | 1.6 (0.7, 3.2) | 52,360 (46,862, 60,133) | 60.2 (54.6, 67.2) |
| 50-75 | 75-100 | 227907 | 14.9% | 66.4% | 33.6% | 54.9% | 46 (35, 55) | 1.5 (0.7, 3.0) | 54,332 (49,961, 60,083) | 60.9 (55.5, 66.9) |
| 25-50 | 0-25 | 29006 | 1.9% | 66.6% | 33.4% | 58.1% | 44 (34, 53) | 1.5 (0.7, 2.9) | 49,759 (44,209, 58,722) | 38.6 (32.7, 44.9) |
| 25-50 | 25-50 | 68794 | 4.5% | 65.4% | 34.6% | 57.2% | 43 (34, 53) | 1.4 (0.7, 2.9) | 63,583 (55,155, 74,153) | 40.0 (33.1, 45.5) |
| 25-50 | 50-75 | 176075 | 11.5% | 64.9% | 35.1% | 56.3% | 44 (34, 54) | 1.5 (0.7, 3.0) | 75,107 (64,092, 92,331) | 37.5 (31.6, 43.7) |
| **25-50** | **75-100** | **284042** | **18.5%** | **65.1%** | **34.9%** | **55.7%** | **45 (34, 54)** | **1.4 (0.6, 2.9)** | **75,830 (66,189, 89,491)** | **37.7 (31.0, 43.4)** |
| 0-25 | 0-25 | 11611 | 0.8% | 64.1% | 35.9% | 58.2% | 45 (35, 53) | 1.3 (0.6, 2.8) | 70,444 (58,651, 90,583) | 16.7 (11.1, 20.8) |
| 0-25 | 25-50 | 46561 | 3.0% | 64.2% | 35.8% | 56.4% | 43 (34, 53) | 1.4 (0.6, 2.8) | 98,034 (79,027, 130,230) | 16.3 (11.3, 20.5) |
| 0-25 | 50-75 | 119064 | 7.8% | 63.4% | 36.6% | 55.8% | 44 (34, 54) | 1.4 (0.7, 2.9) | 123,766 (101,180, 179,109) | 15.1 (9.2, 20.3) |
| 0-25 | 75-100 | 146789 | 9.6% | 64.3% | 35.7% | 55.8% | 46 (35, 55) | 1.4 (0.7, 2.9) | 125,961 (99,965, 174,100) | 16.7 (11.1, 20.9) |
