## Supplement 3 - HCP Count for "Geographic variation in the treatment of spinal disorders: association with health care professional availability, and population socioeconomic status, race, and ethnicity. A retrospective cohort study"

### Supplement 3 - Number of health care providers per 1000 population - Median (Q1, Q3)

| ADI | % NHW | All | | | | Primary Care | | | | Non-Prescriber | | |
|---|---|---|---|---|---|---|---|---|---|---|---|---|
| | | Primary Care | Non-Prescriber | Specialist | Emergency/Urgent | PCP | Nurse | PA | DO | DC | PT | LAc |
| Total | | 1.06 (0.42, 2.38) | 0.45 (0.18, 0.88) | 0.25 (0.03, 1.01) | 0.10 (0.00, 0.31) | 0.67 (0.29, 1.36) | 0.30 (0.09, 0.76) | 0.09 (0.00, 0.28) | 0.00 (0.00, 0.00) | 0.24 (0.10, 0.45) | 0.17 (0.04, 0.42) | 0.00 (0.00, 0.03) |
| 75-100 | All | 1.02 (0.34, 2.32) | 0.17 (0.00, 0.39) | 0.08 (0.00, 0.56) | 0.06 (0.00, 0.29) | 0.65 (0.26, 1.30) | 0.40 (0.12, 0.92) | 0.05 (0.00, 0.22) | 0.00 (0.00, 0.00) | 0.10 (0.00, 0.23) | 0.04 (0.00, 0.20) | 0.00 (0.00, 0.00) |
| 50-75 | | 1.10 (0.41, 2.45) | 0.34 (0.12, 0.68) | 0.20 (0.00, 0.90) | 0.10 (0.00, 0.33) | 0.69 (0.27, 1.36) | 0.37 (0.12, 0.88) | 0.09 (0.00, 0.28) | 0.00 (0.00, 0.00) | 0.20 (0.08, 0.38) | 0.12 (0.00, 0.30) | 0.00 (0.00, 0.00) |
| 25-50 | | 1.08 (0.45, 2.37) | 0.52 (0.25, 0.96) | 0.29 (0.05, 1.03) | 0.10 (0.02, 0.28) | 0.67 (0.31, 1.35) | 0.30 (0.10, 0.74) | 0.11 (0.00, 0.33) | 0.00 (0.00, 0.00) | 0.28 (0.13, 0.49) | 0.21 (0.07, 0.46) | 0.00 (0.00, 0.03) |
| 0-25 | | 0.98 (0.41, 2.32) | 0.68 (0.31, 1.18) | 0.39 (0.08, 1.31) | 0.09 (0.02, 0.32) | 0.66 (0.29, 1.42) | 0.20 (0.04, 0.57) | 0.07 (0.00, 0.24) | 0.00 (0.00, 0.00) | 0.28 (0.13, 0.51) | 0.28 (0.10, 0.59) | 0.02 (0.00, 0.08) |
| All | 0-25 | 0.59 (0.25, 1.34) | 0.14 (0.04, 0.30) | 0.11 (0.01, 0.49) | 0.04 (0.00, 0.14) | 0.36 (0.16, 0.84) | 0.14 (0.04, 0.39) | 0.03 (0.00, 0.11) | 0.00 (0.00, 0.00) | 0.07 (0.01, 0.14) | 0.05 (0.00, 0.15) | 0.00 (0.00, 0.01) |
| | 25-50 | 0.87 (0.39, 1.87) | 0.33 (0.15, 0.59) | 0.25 (0.06, 0.99) | 0.08 (0.02, 0.23) | 0.54 (0.24, 1.13) | 0.20 (0.07, 0.52) | 0.06 (0.00, 0.18) | 0.00 (0.00, 0.00) | 0.16 (0.07, 0.29) | 0.13 (0.03, 0.28) | 0.00 (0.00, 0.04) |
| | 50-75 | 1.17 (0.51, 2.51) | 0.50 (0.23, 0.94) | 0.39 (0.07, 1.34) | 0.11 (0.03, 0.34) | 0.74 (0.32, 1.45) | 0.30 (0.10, 0.76) | 0.11 (0.01, 0.30) | 0.00 (0.00, 0.02) | 0.24 (0.12, 0.44) | 0.21 (0.07, 0.47) | 0.00 (0.00, 0.04) |
| | 75-100 | 1.20 (0.44, 2.61) | 0.55 (0.22, 1.00) | 0.23 (0.00, 0.90) | 0.11 (0.00, 0.35) | 0.74 (0.33, 1.44) | 0.38 (0.13, 0.92) | 0.11 (0.00, 0.35) | 0.00 (0.00, 0.00) | 0.31 (0.15, 0.53) | 0.21 (0.05, 0.49) | 0.00 (0.00, 0.00) |
| 75-100 | 0-25 | 0.66 (0.31, 1.43) | 0.10 (0.00, 0.21) | 0.06 (0.00, 0.35) | 0.03 (0.00, 0.14) | 0.36 (0.17, 0.79) | 0.19 (0.08, 0.48) | 0.05 (0.00, 0.18) | 0.00 (0.00, 0.00) | 0.04 (0.00, 0.10) | 0.03 (0.00, 0.11) | 0.00 (0.00, 0.00) |
| | 25-50 | 1.18 (0.57, 2.87) | 0.23 (0.06, 0.45) | 0.13 (0.00, 1.00) | 0.10 (0.00, 0.38) | 0.70 (0.34, 1.56) | 0.42 (0.13, 1.15) | 0.08 (0.00, 0.30) | 0.00 (0.00, 0.00) | 0.11 (0.00, 0.22) | 0.07 (0.00, 0.24) | 0.00 (0.00, 0.00) |
| | 50-75 | 1.29 (0.38, 2.49) | 0.20 (0.00, 0.40) | 0.13 (0.00, 0.99) | 0.11 (0.00, 0.33) | 0.79 (0.30, 1.40) | 0.46 (0.14, 0.93) | 0.06 (0.00, 0.24) | 0.00 (0.00, 0.00) | 0.12 (0.00, 0.24) | 0.07 (0.00, 0.19) | 0.00 (0.00, 0.00) |
| | 75-100 | 1.30 (0.27, 2.57) | 0.23 (0.00, 0.56) | 0.00 (0.00, 0.40) | 0.06 (0.00, 0.39) | 0.87 (0.38, 1.40) | 0.59 (0.20, 1.20) | 0.00 (0.00, 0.24) | 0.00 (0.00, 0.00) | 0.19 (0.00, 0.40) | 0.07 (0.00, 0.27) | 0.00 (0.00, 0.00) |
| 50-75 | 0-25 | 0.60 (0.24, 1.43) | 0.12 (0.03, 0.26) | 0.08 (0.01, 0.50) | 0.05 (0.00, 0.14) | 0.36 (0.15, 0.95) | 0.15 (0.06, 0.41) | 0.03 (0.00, 0.10) | 0.00 (0.00, 0.00) | 0.07 (0.01, 0.13) | 0.04 (0.00, 0.12) | 0.00 (0.00, 0.00) |
| | 25-50 | 0.89 (0.42, 1.82) | 0.31 (0.14, 0.53) | 0.22 (0.05, 0.87) | 0.08 (0.02, 0.22) | 0.53 (0.24, 1.04) | 0.24 (0.09, 0.55) | 0.09 (0.02, 0.19) | 0.00 (0.00, 0.02) | 0.16 (0.07, 0.28) | 0.10 (0.02, 0.24) | 0.00 (0.00, 0.00) |
| | 50-75 | 1.26 (0.53, 2.70) | 0.39 (0.18, 0.75) | 0.32 (0.04, 1.24) | 0.14 (0.03, 0.36) | 0.73 (0.30, 1.36) | 0.41 (0.14, 0.92) | 0.11 (0.00, 0.34) | 0.00 (0.00, 0.02) | 0.20 (0.10, 0.37) | 0.15 (0.04, 0.35) | 0.00 (0.00, 0.00) |
| | 75-100 | 1.29 (0.41, 2.75) | 0.42 (0.12, 0.79) | 0.15 (0.00, 0.80) | 0.13 (0.00, 0.42) | 0.82 (0.33, 1.52) | 0.50 (0.17, 1.04) | 0.11 (0.00, 0.33) | 0.00 (0.00, 0.00) | 0.27 (0.12, 0.48) | 0.14 (0.00, 0.36) | 0.00 (0.00, 0.00) |
| 25-50 | 0-25 | 0.69 (0.26, 1.21) | 0.20 (0.09, 0.44) | 0.25 (0.05, 0.66) | 0.07 (0.03, 0.16) | 0.47 (0.20, 0.86) | 0.11 (0.03, 0.28) | 0.03 (0.00, 0.09) | 0.00 (0.00, 0.00) | 0.08 (0.04, 0.15) | 0.08 (0.02, 0.22) | 0.00 (0.00, 0.03) |
| | 25-50 | 0.83 (0.37, 1.80) | 0.33 (0.20, 0.59) | 0.24 (0.07, 1.00) | 0.07 (0.02, 0.21) | 0.49 (0.22, 1.02) | 0.18 (0.07, 0.50) | 0.05 (0.00, 0.18) | 0.00 (0.00, 0.00) | 0.16 (0.08, 0.30) | 0.14 (0.06, 0.26) | 0.00 (0.00, 0.04) |
| | 50-75 | 1.16 (0.51, 2.50) | 0.53 (0.28, 0.96) | 0.38 (0.09, 1.26) | 0.11 (0.03, 0.29) | 0.73 (0.30, 1.49) | 0.30 (0.11, 0.74) | 0.12 (0.03, 0.31) | 0.00 (0.00, 0.02) | 0.27 (0.15, 0.46) | 0.21 (0.08, 0.46) | 0.00 (0.00, 0.05) |
| | **75-100 - ref** | **1.19 (0.52, 2.54)** | **0.64 (0.30, 1.09)** | **0.25 (0.00, 0.94)** | **0.11 (0.00, 0.32)** | **0.71 (0.36, 1.35)** | **0.35 (0.13, 0.86)** | **0.14 (0.00, 0.41)** | **0.00 (0.00, 0.00)** | **0.36 (0.19, 0.56)** | **0.25 (0.08, 0.56)** | **0.00 (0.00, 0.02)** |
| 0-25 | 0-25 | 0.38 (0.16, 0.63) | 0.27 (0.09, 0.61) | 0.12 (0.04, 0.39) | 0.02 (0.00, 0.13) | 0.28 (0.15, 0.55) | 0.05 (0.00, 0.15) | 0.00 (0.00, 0.03) | 0.00 (0.00, 0.00) | 0.10 (0.02, 0.23) | 0.10 (0.03, 0.21) | 0.02 (0.00, 0.12) |
| | 25-50 | 0.74 (0.34, 1.70) | 0.41 (0.20, 0.93) | 0.35 (0.10, 1.16) | 0.07 (0.02, 0.23) | 0.55 (0.25, 1.17) | 0.13 (0.02, 0.36) | 0.03 (0.00, 0.12) | 0.00 (0.00, 0.00) | 0.17 (0.08, 0.34) | 0.20 (0.06, 0.42) | 0.04 (0.00, 0.10) |
| | 50-75 | 1.07 (0.48, 2.35) | 0.75 (0.36, 1.20) | 0.59 (0.16, 1.55) | 0.11 (0.03, 0.39) | 0.77 (0.35, 1.50) | 0.20 (0.05, 0.51) | 0.08 (0.00, 0.24) | 0.00 (0.00, 0.01) | 0.29 (0.14, 0.53) | 0.33 (0.13, 0.61) | 0.03 (0.00, 0.10) |
| | 75-100 | 1.04 (0.43, 2.58) | 0.74 (0.35, 1.25) | 0.30 (0.04, 1.21) | 0.10 (0.00, 0.28) | 0.67 (0.29, 1.48) | 0.25 (0.08, 0.71) | 0.09 (0.00, 0.31) | 0.00 (0.00, 0.00) | 0.33 (0.17, 0.57) | 0.30 (0.12, 0.65) | 0.00 (0.00, 0.06) |

| ADI | % NHW | Specialist | | | | | | | Emergency/Urgent Care | | |
|---|---|---|---|---|---|---|---|---|---|---|---|
| | | OS | PMR | PM | NS | Neuro | Rheum | MD (Oth) | EM | Rad | UC |
| Total | | 0.02 (0.00, 0.13) | 0.00 (0.00, 0.05) | 0.00 (0.00, 0.03) | 0.00 (0.00, 0.00) | 0.00 (0.00, 0.05) | 0.00 (0.00, 0.00) | 0.18 (0.01, 0.74) | 0.04 (0.00, 0.15) | 0.02 (0.00, 0.11) | 0.00 (0.00, 0.03) |
| 75-100 | All | 0.00 (0.00, 0.09) | 0.00 (0.00, 0.02) | 0.00 (0.00, 0.00) | 0.00 (0.00, 0.00) | 0.00 (0.00, 0.02) | 0.00 (0.00, 0.00) | 0.07 (0.00, 0.45) | 0.03 (0.00, 0.19) | 0.00 (0.00, 0.10) | 0.00 (0.00, 0.00) |
| 50-75 | | 0.00 (0.00, 0.13) | 0.00 (0.00, 0.04) | 0.00 (0.00, 0.03) | 0.00 (0.00, 0.00) | 0.00 (0.00, 0.04) | 0.00 (0.00, 0.00) | 0.16 (0.00, 0.65) | 0.05 (0.00, 0.18) | 0.02 (0.00, 0.12) | 0.00 (0.00, 0.02) |
| 25-50 | | 0.03 (0.00, 0.13) | 0.00 (0.00, 0.05) | 0.00 (0.00, 0.04) | 0.00 (0.00, 0.00) | 0.00 (0.00, 0.05) | 0.00 (0.00, 0.02) | 0.19 (0.03, 0.75) | 0.04 (0.00, 0.13) | 0.03 (0.00, 0.11) | 0.00 (0.00, 0.03) |
| 0-25 | | 0.03 (0.00, 0.16) | 0.00 (0.00, 0.07) | 0.00 (0.00, 0.04) | 0.00 (0.00, 0.00) | 0.00 (0.00, 0.07) | 0.00 (0.00, 0.03) | 0.26 (0.05, 0.95) | 0.03 (0.00, 0.11) | 0.02 (0.00, 0.13) | 0.00 (0.00, 0.03) |
| All | 0-25 | 0.00 (0.00, 0.05) | 0.00 (0.00, 0.02) | 0.00 (0.00, 0.02) | 0.00 (0.00, 0.00) | 0.00 (0.00, 0.03) | 0.00 (0.00, 0.00) | 0.08 (0.00, 0.35) | 0.01 (0.00, 0.06) | 0.00 (0.00, 0.07) | 0.00 (0.00, 0.01) |
| | 25-50 | 0.02 (0.00, 0.12) | 0.00 (0.00, 0.04) | 0.00 (0.00, 0.03) | 0.00 (0.00, 0.00) | 0.00 (0.00, 0.05) | 0.00 (0.00, 0.02) | 0.18 (0.03, 0.67) | 0.03 (0.00, 0.10) | 0.02 (0.00, 0.09) | 0.00 (0.00, 0.03) |
| | 50-75 | 0.04 (0.00, 0.16) | 0.00 (0.00, 0.06) | 0.00 (0.00, 0.04) | 0.00 (0.00, 0.01) | 0.00 (0.00, 0.07) | 0.00 (0.00, 0.02) | 0.28 (0.05, 0.93) | 0.04 (0.00, 0.15) | 0.03 (0.00, 0.14) | 0.00 (0.00, 0.03) |
| | 75-100 | 0.02 (0.00, 0.15) | 0.00 (0.00, 0.04) | 0.00 (0.00, 0.03) | 0.00 (0.00, 0.00) | 0.00 (0.00, 0.04) | 0.00 (0.00, 0.00) | 0.17 (0.00, 0.68) | 0.05 (0.00, 0.19) | 0.00 (0.00, 0.12) | 0.00 (0.00, 0.03) |
| 75-100 | 0-25 | 0.00 (0.00, 0.03) | 0.00 (0.00, 0.02) | 0.00 (0.00, 0.00) | 0.00 (0.00, 0.00) | 0.00 (0.00, 0.02) | 0.00 (0.00, 0.00) | 0.05 (0.00, 0.27) | 0.00 (0.00, 0.06) | 0.00 (0.00, 0.06) | 0.00 (0.00, 0.00) |
| | 25-50 | 0.00 (0.00, 0.15) | 0.00 (0.00, 0.05) | 0.00 (0.00, 0.02) | 0.00 (0.00, 0.00) | 0.00 (0.00, 0.05) | 0.00 (0.00, 0.00) | 0.11 (0.00, 0.84) | 0.05 (0.00, 0.20) | 0.00 (0.00, 0.14) | 0.00 (0.00, 0.00) |
| | 50-75 | 0.00 (0.00, 0.12) | 0.00 (0.00, 0.03) | 0.00 (0.00, 0.00) | 0.00 (0.00, 0.00) | 0.00 (0.00, 0.04) | 0.00 (0.00, 0.00) | 0.09 (0.00, 0.79) | 0.07 (0.00, 0.20) | 0.00 (0.00, 0.12) | 0.00 (0.00, 0.00) |
| | 75-100 | 0.00 (0.00, 0.10) | 0.00 (0.00, 0.00) | 0.00 (0.00, 0.00) | 0.00 (0.00, 0.00) | 0.00 (0.00, 0.00) | 0.00 (0.00, 0.00) | 0.06 (0.00, 0.35) | 0.09 (0.00, 0.30) | 0.00 (0.00, 0.11) | 0.00 (0.00, 0.00) |
| 50-75 | 0-25 | 0.00 (0.00, 0.04) | 0.00 (0.00, 0.02) | 0.00 (0.00, 0.02) | 0.00 (0.00, 0.00) | 0.00 (0.00, 0.02) | 0.00 (0.00, 0.00) | 0.06 (0.00, 0.39) | 0.02 (0.00, 0.05) | 0.00 (0.00, 0.07) | 0.00 (0.00, 0.01) |
| | 25-50 | 0.02 (0.00, 0.11) | 0.00 (0.00, 0.04) | 0.00 (0.00, 0.03) | 0.00 (0.00, 0.00) | 0.00 (0.00, 0.04) | 0.00 (0.00, 0.00) | 0.15 (0.02, 0.59) | 0.03 (0.00, 0.12) | 0.02 (0.00, 0.08) | 0.00 (0.00, 0.02) |
| | 50-75 | 0.03 (0.00, 0.16) | 0.00 (0.00, 0.04) | 0.00 (0.00, 0.04) | 0.00 (0.00, 0.00) | 0.00 (0.00, 0.06) | 0.00 (0.00, 0.00) | 0.22 (0.03, 0.86) | 0.05 (0.00, 0.18) | 0.04 (0.00, 0.14) | 0.00 (0.00, 0.03) |
| | 75-100 | 0.00 (0.00, 0.14) | 0.00 (0.00, 0.03) | 0.00 (0.00, 0.02) | 0.00 (0.00, 0.00) | 0.00 (0.00, 0.04) | 0.00 (0.00, 0.00) | 0.16 (0.00, 0.64) | 0.09 (0.00, 0.27) | 0.00 (0.00, 0.12) | 0.00 (0.00, 0.03) |
| 25-50 | 0-25 | 0.02 (0.00, 0.06) | 0.00 (0.00, 0.03) | 0.00 (0.00, 0.02) | 0.00 (0.00, 0.00) | 0.00 (0.00, 0.04) | 0.00 (0.00, 0.01) | 0.14 (0.03, 0.50) | 0.02 (0.00, 0.06) | 0.02 (0.00, 0.08) | 0.00 (0.00, 0.02) |
| | 25-50 | 0.02 (0.00, 0.11) | 0.00 (0.00, 0.04) | 0.00 (0.00, 0.03) | 0.00 (0.00, 0.00) | 0.00 (0.00, 0.05) | 0.00 (0.00, 0.02) | 0.17 (0.04, 0.66) | 0.02 (0.00, 0.09) | 0.02 (0.00, 0.08) | 0.00 (0.00, 0.03) |
| | 50-75 | 0.03 (0.00, 0.15) | 0.01 (0.00, 0.06) | 0.00 (0.00, 0.05) | 0.00 (0.00, 0.01) | 0.00 (0.00, 0.07) | 0.00 (0.00, 0.02) | 0.28 (0.05, 0.89) | 0.04 (0.00, 0.13) | 0.03 (0.00, 0.11) | 0.00 (0.00, 0.04) |
| | **75-100 - ref** | **0.03 (0.00, 0.15)** | **0.00 (0.00, 0.05)** | **0.00 (0.00, 0.03)** | **0.00 (0.00, 0.00)** | **0.00 (0.00, 0.04)** | **0.00 (0.00, 0.00)** | **0.17 (0.00, 0.71)** | **0.04 (0.00, 0.16)** | **0.02 (0.00, 0.12)** | **0.00 (0.00, 0.04)** |
| 0-25 | 0-25 | 0.00 (0.00, 0.05) | 0.00 (0.00, 0.03) | 0.00 (0.00, 0.00) | 0.00 (0.00, 0.00) | 0.00 (0.00, 0.00) | 0.00 (0.00, 0.00) | 0.11 (0.03, 0.23) | 0.00 (0.00, 0.04) | 0.00 (0.00, 0.05) | 0.00 (0.00, 0.00) |
| | 25-50 | 0.03 (0.00, 0.14) | 0.00 (0.00, 0.06) | 0.00 (0.00, 0.03) | 0.00 (0.00, 0.02) | 0.00 (0.00, 0.07) | 0.00 (0.00, 0.02) | 0.23 (0.05, 0.80) | 0.02 (0.00, 0.06) | 0.03 (0.00, 0.11) | 0.00 (0.00, 0.03) |
| | 50-75 | 0.05 (0.00, 0.18) | 0.03 (0.00, 0.08) | 0.00 (0.00, 0.04) | 0.00 (0.00, 0.02) | 0.02 (0.00, 0.09) | 0.00 (0.00, 0.03) | 0.39 (0.09, 1.13) | 0.03 (0.00, 0.14) | 0.04 (0.00, 0.17) | 0.00 (0.00, 0.04) |
| | 75-100 | 0.02 (0.00, 0.17) | 0.00 (0.00, 0.06) | 0.00 (0.00, 0.03) | 0.00 (0.00, 0.00) | 0.00 (0.00, 0.07) | 0.00 (0.00, 0.00) | 0.20 (0.00, 0.84) | 0.04 (0.00, 0.11) | 0.00 (0.00, 0.12) | 0.00 (0.00, 0.02) |

ADI=Area Deprivation Index, NHW=Non-Hispanic White, PCP=Primary Care Provider, PA=Physician Assistant, DO=Doctor of Osteopathy, DC=Doctor of Chiropractic, PT=Physical Therapist, LAc=Licensed Acupuncturist, OS=Orthpedic Surgeon, PMR=Physical Medicine & Rehabilitation, PM=Pain Management, NS=Neurosurgeon, Neuro=Neurologist, Rheum=Rheumatologist, MD Oth=Other MD specialty, EM=Emergency Medicine, Rad=Radiologist, UC=Urgent Care

Cells with red text denote that the effect of provider type on service usage was found not to be significantly different from that of ADI 0-25 % NHW 75-100 reference (Mann-Whitney U p > 0.001)

Cells with black text denote that the effect of provider type on service usage was found to be significantly different from that of ADI 0-25 % NHW 75-100 reference (Mann-Whitney U p > 0.001)
