## Supplement 3a - HCP Count Figure for "Geographic variation in the treatment of spinal disorders: association with health care professional availability, and population socioeconomic status, race, and ethnicity. A retrospective cohort study"

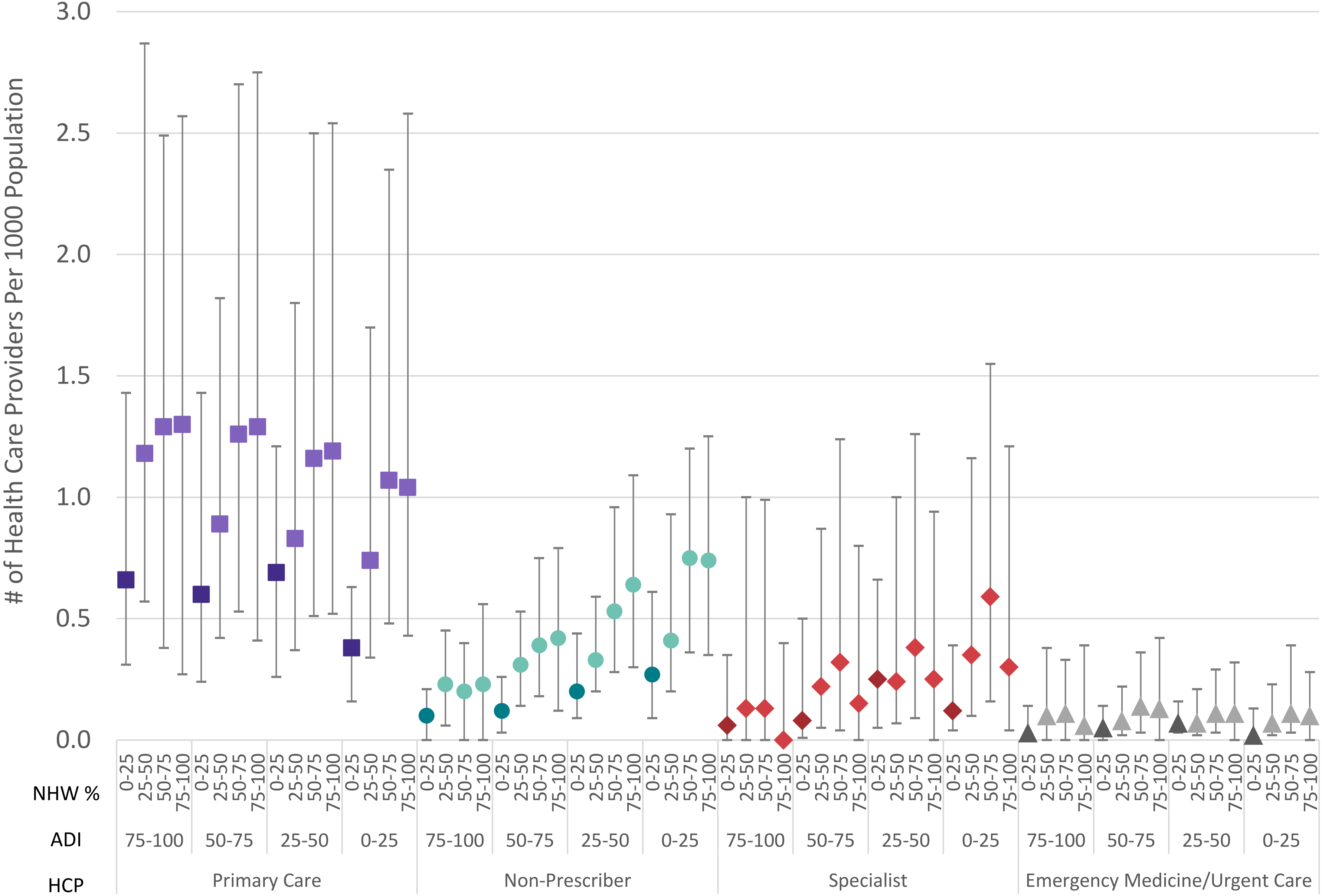


**Supplement 3a** - Median and interquartile range (IQR) for number of health care providers per 1000 population in 5 digit zip code by zip code Area Deprivation Index (ADI) and percent non-Hispanic white population (NHW%).
