## Supplement 4 - Initial Contact HCP for "Geographic variation in the treatment of spinal disorders: association with health care professional availability, and population socioeconomic status, race, and ethnicity. A retrospective cohort study"

### Supplement 4 - % of spinal disorder episodes where individual initially contacted health care provider type

| ADI | % NHW | Episodes | All | | | | Primary Care | | | | Non-Prescriber | | | Specialist | | | | | | | Emergency/ Urgent Care | | |
|---|---|---|---|---|---|---|---|---|---|---|---|---|---|---|---|---|---|---|---|---|---|---|---|
| | | | Primary Care | Non-Prescriber | Specialist | Emergency/ Urgent | PCP | Nurse | PA | DO | DC | PT | LAc | OS | PMR | PM | NS | Neuro | Rheu | MD (Oth) | EM | Rad | UC |
| Total | | 1510869 | 43.8% | 35.2% | 15.0% | 6.0% | 32.8% | 6.5% | 4.3% | 0.1% | 33.5% | 0.9% | 0.7% | 6.0% | 2.5% | 1.1% | 1.1% | 1.3% | 0.9% | 2.0% | 2.9% | 2.4% | 0.5% |
| 75-100 | All | 128401 | 54.9% | 25.6% | 11.7% | 7.8% | 38.4% | 11.8% | 4.6% | 0.0% | 24.9% | 0.4% | 0.1% | 4.4% | 1.6% | 0.9% | 1.1% | 1.1% | 0.9% | 1.7% | 4.3% | 3.1% | 0.4% |
| 50-75 | | 500526 | 47.1% | 34.0% | 12.7% | 6.2% | 34.1% | 8.1% | 4.8% | 0.1% | 33.2% | 0.7% | 0.3% | 4.9% | 1.9% | 1.1% | 1.1% | 1.2% | 0.8% | 1.9% | 3.3% | 2.4% | 0.4% |
| 25-50 | | 557917 | 41.8% | 37.7% | 14.8% | 5.7% | 31.5% | 5.5% | 4.5% | 0.1% | 36.1% | 1.1% | 0.7% | 5.9% | 2.5% | 1.1% | 1.1% | 1.3% | 0.9% | 2.0% | 2.8% | 2.2% | 0.7% |
| 0-25 | | 324025 | 37.8% | 36.7% | 20.0% | 5.6% | 31.0% | 3.3% | 3.2% | 0.1% | 33.3% | 1.7% | 1.7% | 8.6% | 4.2% | 1.0% | 1.1% | 1.8% | 1.0% | 2.4% | 2.4% | 2.4% | 0.8% |
| All | 0-25 | 125079 | 53.9% | 20.5% | 16.3% | 9.3% | 43.9% | 6.0% | 4.0% | 0.0% | 18.6% | 0.9% | 1.0% | 6.0% | 2.5% | 1.2% | 1.0% | 1.9% | 1.2% | 2.4% | 5.3% | 3.1% | 0.9% |
| | 25-50 | 223789 | 46.9% | 29.1% | 17.3% | 6.7% | 37.4% | 5.5% | 3.9% | 0.1% | 27.0% | 1.0% | 1.0% | 7.4% | 2.9% | 1.2% | 1.0% | 1.7% | 1.0% | 2.2% | 3.5% | 2.5% | 0.7% |
| | 50-75 | 461777 | 43.9% | 33.7% | 16.7% | 5.7% | 33.7% | 5.9% | 4.2% | 0.1% | 31.8% | 1.1% | 0.8% | 6.9% | 2.8% | 1.2% | 1.1% | 1.5% | 0.9% | 2.2% | 2.8% | 2.3% | 0.5% |
| | 75-100 | 700224 | 41.3% | 40.4% | 12.9% | 5.4% | 29.2% | 7.2% | 4.7% | 0.1% | 39.1% | 0.9% | 0.4% | 5.0% | 2.3% | 0.9% | 1.0% | 1.2% | 0.8% | 1.8% | 2.6% | 2.3% | 0.5% |
| 75-100 | 0-25 | 35387 | 59.3% | 16.9% | 13.1% | 10.7% | 44.4% | 9.1% | 5.7% | 0.0% | 16.1% | 0.6% | 0.3% | 4.9% | 1.9% | 1.0% | 1.1% | 1.3% | 1.0% | 1.9% | 6.4% | 3.4% | 0.7% |
| | 25-50 | 24343 | 55.5% | 23.1% | 13.0% | 8.4% | 38.5% | 12.0% | 4.9% | 0.0% | 22.3% | 0.5% | 0.1% | 5.0% | 1.8% | 1.2% | 0.9% | 1.2% | 0.9% | 1.9% | 4.6% | 3.4% | 0.4% |
| | 50-75 | 27185 | 55.7% | 25.0% | 12.3% | 7.0% | 39.2% | 12.2% | 4.4% | 0.1% | 24.5% | 0.5% | 0.1% | 4.4% | 1.5% | 1.2% | 1.3% | 1.2% | 0.9% | 1.7% | 4.0% | 2.8% | 0.3% |
| | 75-100 | 41486 | 50.6% | 34.1% | 9.7% | 5.6% | 33.1% | 13.6% | 3.9% | 0.0% | 33.7% | 0.4% | 0.0% | 3.6% | 1.2% | 0.6% | 1.0% | 0.8% | 0.8% | 1.6% | 2.7% | 2.7% | 0.1% |
| 50-75 | 0-25 | 49075 | 54.3% | 20.6% | 16.0% | 9.1% | 44.4% | 5.7% | 4.0% | 0.1% | 19.3% | 0.9% | 0.6% | 6.2% | 2.5% | 1.2% | 0.9% | 1.9% | 1.0% | 2.4% | 5.4% | 3.2% | 0.6% |
| | 25-50 | 84091 | 49.7% | 27.3% | 16.1% | 6.9% | 38.3% | 6.3% | 5.0% | 0.1% | 26.2% | 0.8% | 0.4% | 6.7% | 2.5% | 1.2% | 1.1% | 1.7% | 1.0% | 2.1% | 3.9% | 2.5% | 0.6% |
| | 50-75 | 139453 | 48.5% | 31.3% | 14.0% | 6.2% | 35.6% | 7.9% | 5.0% | 0.1% | 30.3% | 0.7% | 0.3% | 5.5% | 2.0% | 1.2% | 1.2% | 1.3% | 0.9% | 1.8% | 3.2% | 2.5% | 0.5% |
| | 75-100 | 227907 | 43.9% | 40.5% | 10.2% | 5.4% | 29.8% | 9.2% | 4.7% | 0.1% | 39.9% | 0.5% | 0.1% | 3.7% | 1.5% | 0.8% | 0.9% | 1.0% | 0.6% | 1.5% | 2.7% | 2.3% | 0.4% |
| 25-50 | 0-25 | 29006 | 49.7% | 22.2% | 19.9% | 8.2% | 43.2% | 3.8% | 2.7% | 0.0% | 19.5% | 1.1% | 1.7% | 7.1% | 2.9% | 1.5% | 0.9% | 2.9% | 1.8% | 2.9% | 4.2% | 2.7% | 1.2% |
| | 25-50 | 68794 | 44.8% | 31.4% | 17.6% | 6.2% | 36.9% | 4.3% | 3.6% | 0.1% | 29.5% | 1.1% | 1.0% | 7.5% | 3.0% | 1.2% | 1.0% | 1.5% | 1.0% | 2.2% | 3.2% | 2.2% | 0.8% |
| | 50-75 | 176075 | 42.8% | 35.3% | 16.6% | 5.2% | 33.2% | 5.4% | 4.3% | 0.1% | 33.7% | 0.9% | 0.7% | 6.8% | 2.8% | 1.3% | 1.2% | 1.5% | 0.9% | 2.1% | 2.5% | 2.1% | 0.5% |
| | **75-100 ref** | **284042** | **39.9%** | **42.1%** | **12.6%** | **5.5%** | **28.6%** | **6.3%** | **5.0%** | **0.1%** | **40.6%** | **1.0%** | **0.4%** | **4.9%** | **2.2%** | **0.9%** | **1.0%** | **1.0%** | **0.8%** | **1.8%** | **2.6%** | **2.3%** | **0.6%** |
| 0-25 | 0-25 | 11611 | 45.3% | 27.4% | 18.7% | 8.7% | 41.3% | 2.0% | 1.8% | 0.2% | 21.3% | 1.7% | 4.4% | 7.5% | 3.5% | 0.9% | 0.8% | 2.4% | 1.1% | 2.4% | 3.8% | 3.4% | 1.7% |
| | 25-50 | 46561 | 39.9% | 32.4% | 21.7% | 6.0% | 36.0% | 2.2% | 1.7% | 0.0% | 28.2% | 1.6% | 2.6% | 9.5% | 4.5% | 1.0% | 1.0% | 2.2% | 1.2% | 2.6% | 2.4% | 2.4% | 1.2% |
| | 50-75 | 119064 | 36.8% | 36.6% | 21.2% | 5.4% | 31.3% | 2.7% | 2.8% | 0.1% | 33.0% | 1.8% | 2.0% | 9.3% | 4.4% | 1.1% | 1.0% | 1.8% | 1.0% | 2.5% | 2.3% | 2.3% | 0.8% |
| | 75-100 | 146789 | 37.3% | 38.7% | 18.7% | 5.3% | 28.8% | 4.3% | 4.0% | 0.1% | 36.0% | 1.6% | 1.1% | 8.1% | 3.9% | 0.9% | 1.2% | 1.6% | 0.9% | 2.2% | 2.3% | 2.4% | 0.5% |

PCP=Primary Care Provider, PA=Physician Assistant, DO=Doctor of Osteopathy, DC=Doctor of Chiropractic, PT=Physical Therapist, LAc=Licensed Acupuncturist, OS=Orthpedic Surgeon, PMR=Physical Medicine & Rehabilitation, PM=Pain Management, NS=Neurosurgeon, Neuro=Neurologist, Rheum=Rheumatologist, MD Oth=Other MD specialty, EM=Emergency Medicine, Rad=Radiologist, UC=Urgent Care

Cells with red text denote that the effect of provider type on service usage was found not to be significantly different from that of ADI 0-25 % NHW 75-100 reference (Fisher's Exact $p > 0.001$)
Cells with black text denote that the effect of provider type on service usage was found to be significantly different from that of ADI 0-25 % NHW 75-100 reference (Fisher's Exact $p < 0.001$)
