## Supplement 4a - Initial Contact HCP Risk Ratio for "Geographic variation in the treatment of spinal disorders: association with health care professional availability, and population socioeconomic status, race, and ethnicity. A retrospective cohort study"

**Supplement 4a - Risk ratio and 95% confidence interval for % of spinal episodes by initial contact health care provider compared to reference segment of 25-50 Area Deprivation Index (ADI) and 75-100% non-Hispanic white (NHW) population**

| ADI | % NHW | All | | | | Primary Care (PC) | | | | Non-Prescriber (NP) | | | Specialist (Spec) | | | | | | | Emergency Medicine/ Urgent Care (EM/UC) | | |
|---|---|---|---|---|---|---|---|---|---|---|---|---|---|---|---|---|---|---|---|---|---|---|
| | | PC | NP | Spec | EM/UC | PCP | Nurse | PA | DO | DC | PT | LAc | OS | PMR | PM | NS | Neuro | Rheu | MD (Oth) | EM | Rad | UC |
| 75-100 | 0-25 | 1.34 (1.32, 1.35) | 0.36 (0.35, 0.37) | 0.93 (0.90, 0.96) | 1.73 (1.66, 1.80) | 1.40 (1.38, 1.42) | 1.31 (1.25, 1.37) | 1.03 (0.98, 1.09) | 0.26 (0.13, 0.50) | 0.36 (0.34, 0.37) | 0.46 (0.39, 0.56) | 0.63 (0.48, 0.82) | 0.90 (0.84, 0.95) | 0.75 (0.68, 0.82) | 1.05 (0.93, 1.20) | 1.03 (0.91, 1.16) | 1.03 (0.92, 1.16) | 1.20 (1.05, 1.37) | 0.97 (0.88, 1.07) | 2.23 (2.11, 2.36) | 1.35 (1.26, 1.45) | 1.08 (0.93, 1.25) |
| | 25-50 | 1.32 (1.30, 1.34) | 0.52 (0.51, 0.54) | 0.98 (0.94, 1.02) | 1.42 (1.35, 1.50) | 1.28 (1.26, 1.31) | 1.82 (1.75, 1.90) | 0.94 (0.88, 1.00) | 0.21 (0.09, 0.50) | 0.52 (0.51, 0.54) | 0.52 (0.42, 0.63) | 0.46 (0.32, 0.67) | 0.97 (0.91, 1.04) | 0.78 (0.70, 0.87) | 1.24 (1.08, 1.43) | 0.91 (0.78, 1.05) | 1.04 (0.90, 1.19) | 1.19 (1.01, 1.39) | 1.01 (0.91, 1.13) | 1.70 (1.58, 1.82) | 1.37 (1.26, 1.48) | 0.53 (0.41, 0.68) |
| | 50-75 | 1.35 (1.33, 1.38) | 0.58 (0.56, 0.59) | 0.95 (0.91, 0.98) | 1.23 (1.17, 1.30) | 1.33 (1.30, 1.35) | 1.89 (1.81, 1.96) | 0.85 (0.79, 0.91) | 0.60 (0.36, 0.99) | 0.58 (0.57, 0.60) | 0.50 (0.41, 0.60) | 0.26 (0.17, 0.41) | 0.87 (0.82, 0.93) | 0.66 (0.59, 0.74) | 1.33 (1.17, 1.52) | 1.21 (1.07, 1.38) | 1.11 (0.97, 1.26) | 1.21 (1.04, 1.40) | 0.96 (0.86, 1.07) | 1.50 (1.39, 1.61) | 1.17 (1.08, 1.28) | 0.37 (0.28, 0.49) |
| | 75-100 | 1.25 (1.24, 1.27) | 0.80 (0.79, 0.82) | 0.76 (0.73, 0.79) | 1.00 (0.95, 1.05) | 1.15 (1.13, 1.17) | 2.14 (2.07, 2.21) | 0.77 (0.73, 0.81) | 0.49 (0.31, 0.77) | 0.82 (0.81, 0.83) | 0.34 (0.28, 0.42) | 0.04 (0.01, 0.10) | 0.72 (0.68, 0.76) | 0.54 (0.49, 0.60) | 0.76 (0.67, 0.88) | 1.05 (0.94, 1.17) | 0.74 (0.65, 0.84) | 1.02 (0.89, 1.16) | 0.87 (0.79, 0.96) | 1.02 (0.95, 1.10) | 1.17 (1.09, 1.26) | 0.26 (0.20, 0.34) |
| 50-75 | 0-25 | 1.19 (1.17, 1.20) | 0.43 (0.42, 0.44) | 1.11 (1.08, 1.14) | 1.44 (1.38, 1.49) | 1.36 (1.34, 1.38) | 0.81 (0.77, 0.84) | 0.70 (0.66, 0.74) | 0.52 (0.34, 0.78) | 0.41 (0.40, 0.42) | 0.71 (0.62, 0.80) | 1.55 (1.33, 1.81) | 1.08 (1.04, 1.14) | 1.00 (0.93, 1.07) | 1.18 (1.06, 1.32) | 0.75 (0.66, 0.84) | 1.51 (1.39, 1.65) | 1.26 (1.12, 1.41) | 1.18 (1.09, 1.27) | 1.81 (1.72, 1.91) | 1.19 (1.12, 1.27) | 0.79 (0.68, 0.92) |
| | 25-50 | 1.16 (1.15, 1.17) | 0.60 (0.60, 0.61) | 1.19 (1.17, 1.22) | 1.15 (1.11, 1.19) | 1.25 (1.23, 1.27) | 0.95 (0.92, 0.98) | 0.92 (0.89, 0.96) | 0.58 (0.43, 0.79) | 0.60 (0.59, 0.61) | 0.71 (0.64, 0.78) | 0.95 (0.82, 1.11) | 1.27 (1.23, 1.32) | 1.03 (0.98, 1.09) | 1.33 (1.23, 1.45) | 0.98 (0.90, 1.07) | 1.39 (1.29, 1.49) | 1.19 (1.09, 1.31) | 1.09 (1.02, 1.16) | 1.37 (1.31, 1.44) | 0.98 (0.92, 1.03) | 0.85 (0.76, 0.95) |
| | 50-75 | 1.18 (1.17, 1.19) | 0.72 (0.71, 0.73) | 1.08 (1.06, 1.10) | 1.08 (1.05, 1.11) | 1.21 (1.20, 1.22) | 1.23 (1.20, 1.26) | 0.98 (0.95, 1.01) | 0.73 (0.58, 0.91) | 0.72 (0.72, 0.73) | 0.69 (0.63, 0.75) | 0.57 (0.49, 0.66) | 1.08 (1.05, 1.12) | 0.89 (0.85, 0.94) | 1.35 (1.26, 1.45) | 1.12 (1.05, 1.20) | 1.18 (1.11, 1.26) | 1.18 (1.09, 1.28) | 1.06 (1.00, 1.12) | 1.20 (1.15, 1.25) | 1.03 (0.99, 1.08) | 0.74 (0.67, 0.82) |
| | 75-100 | 1.10 (1.10, 1.11) | 0.97 (0.96, 0.98) | 0.81 (0.80, 0.83) | 0.97 (0.95, 1.00) | 1.05 (1.04, 1.06) | 1.48 (1.45, 1.51) | 0.96 (0.94, 0.99) | 0.81 (0.67, 0.98) | 0.98 (0.98, 0.99) | 0.54 (0.50, 0.58) | 0.21 (0.17, 0.25) | 0.76 (0.74, 0.78) | 0.71 (0.68, 0.75) | 0.86 (0.81, 0.93) | 0.94 (0.88, 1.00) | 0.90 (0.85, 0.96) | 0.91 (0.85, 0.98) | 0.88 (0.84, 0.92) | 1.05 (1.01, 1.09) | 0.99 (0.95, 1.03) | 0.55 (0.50, 0.60) |
| 25-50 | 0-25 | 1.06 (1.04, 1.08) | 0.45 (0.44, 0.46) | 1.34 (1.30, 1.38) | 1.24 (1.18, 1.31) | 1.29 (1.26, 1.31) | 0.51 (0.47, 0.55) | 0.46 (0.42, 0.50) | 0.31 (0.16, 0.61) | 0.41 (0.39, 0.42) | 0.91 (0.79, 1.05) | 4.19 (3.68, 4.77) | 1.22 (1.16, 1.29) | 1.08 (0.99, 1.18) | 1.37 (1.21, 1.55) | 0.81 (0.70, 0.94) | 2.26 (2.06, 2.48) | 2.06 (1.83, 2.31) | 1.43 (1.31, 1.56) | 1.39 (1.29, 1.49) | 0.98 (0.90, 1.07) | 1.61 (1.40, 1.85) |
| | 25-50 | 1.04 (1.03, 1.05) | 0.69 (0.68, 0.70) | 1.29 (1.26, 1.32) | 1.04 (1.00, 1.08) | 1.20 (1.18, 1.22) | 0.63 (0.60, 0.66) | 0.66 (0.63, 0.70) | 0.55 (0.39, 0.77) | 0.67 (0.66, 0.68) | 0.97 (0.88, 1.07) | 2.71 (2.42, 3.03) | 1.41 (1.36, 1.46) | 1.27 (1.20, 1.34) | 1.28 (1.17, 1.40) | 0.93 (0.84, 1.02) | 1.37 (1.26, 1.48) | 1.16 (1.05, 1.29) | 1.19 (1.11, 1.27) | 1.12 (1.06, 1.19) | 0.90 (0.84, 0.96) | 1.24 (1.12, 1.39) |
| | 50-75 | 1.03 (1.02, 1.04) | 0.81 (0.80, 0.81) | 1.26 (1.24, 1.28) | 0.90 (0.87, 0.92) | 1.11 (1.10, 1.12) | 0.81 (0.79, 0.83) | 0.83 (0.81, 0.86) | 1.15 (0.96, 1.38) | 0.79 (0.79, 0.80) | 0.92 (0.86, 0.99) | 2.01 (1.83, 2.22) | 1.33 (1.29, 1.37) | 1.22 (1.17, 1.28) | 1.37 (1.28, 1.46) | 1.09 (1.02, 1.16) | 1.36 (1.28, 1.44) | 1.17 (1.09, 1.26) | 1.16 (1.10, 1.22) | 0.94 (0.90, 0.98) | 0.86 (0.82, 0.90) | 0.84 (0.77, 0.92) |
| | 75-100 | reference | | | | | | | | | | | | | | | | | | | | |
| 0-25 | 0-25 | 0.95 (0.92, 0.98) | 0.55 (0.53, 0.57) | 1.24 (1.18, 1.30) | 1.32 (1.22, 1.42) | 1.21 (1.17, 1.25) | 0.27 (0.23, 0.32) | 0.32 (0.27, 0.38) | 0.52 (0.23, 1.18) | 0.44 (0.42, 0.46) | 1.42 (1.19, 1.69) | 11.02 (9.71, 12.51) | 1.27 (1.17, 1.38) | 1.35 (1.20, 1.52) | 0.84 (0.66, 1.07) | 0.57 (0.44, 0.75) | 1.96 (1.69, 2.27) | 1.22 (0.97, 1.52) | 1.15 (0.99, 1.33) | 1.21 (1.08, 1.37) | 1.20 (1.05, 1.35) | 2.17 (1.81, 2.61) |
| | 25-50 | 0.89 (0.88, 0.90) | 0.69 (0.67, 0.70) | 1.54 (1.50, 1.58) | 0.97 (0.92, 1.01) | 1.12 (1.10, 1.14) | 0.30 (0.27, 0.32) | 0.31 (0.29, 0.34) | 0.48 (0.31, 0.74) | 0.62 (0.61, 0.63) | 1.40 (1.28, 1.55) | 6.84 (6.21, 7.54) | 1.72 (1.66, 1.79) | 1.81 (1.71, 1.92) | 0.97 (0.86, 1.09) | 0.85 (0.76, 0.96) | 1.75 (1.60, 1.90) | 1.31 (1.17, 1.47) | 1.34 (1.24, 1.44) | 0.83 (0.77, 0.89) | 0.93 (0.86, 1.00) | 1.68 (1.50, 1.88) |
| | 50-75 | 0.84 (0.83, 0.85) | 0.79 (0.78, 0.80) | 1.52 (1.50, 1.55) | 0.88 (0.85, 0.91) | 0.99 (0.98, 1.00) | 0.39 (0.37, 0.41) | 0.51 (0.48, 0.53) | 0.83 (0.66, 1.05) | 0.73 (0.73, 0.74) | 1.65 (1.55, 1.76) | 5.11 (4.69, 5.57) | 1.70 (1.66, 1.75) | 1.79 (1.72, 1.87) | 1.13 (1.05, 1.23) | 0.92 (0.86, 1.00) | 1.61 (1.51, 1.71) | 1.15 (1.06, 1.25) | 1.33 (1.27, 1.41) | 0.80 (0.76, 0.84) | 0.87 (0.82, 0.92) | 1.21 (1.11, 1.33) |
| | 75-100 | 0.89 (0.88, 0.90) | 0.88 (0.87, 0.88) | 1.42 (1.39, 1.44) | 0.90 (0.87, 0.93) | 0.96 (0.95, 0.97) | 0.66 (0.64, 0.68) | 0.78 (0.75, 0.81) | 1.33 (1.10, 1.59) | 0.84 (0.83, 0.85) | 1.57 (1.48, 1.68) | 2.86 (2.60, 3.13) | 1.56 (1.51, 1.60) | 1.68 (1.62, 1.75) | 1.06 (0.98, 1.14) | 1.09 (1.02, 1.17) | 1.38 (1.30, 1.47) | 1.14 (1.05, 1.23) | 1.20 (1.14, 1.26) | 0.84 (0.81, 0.88) | 0.97 (0.92, 1.02) | 0.85 (0.78, 0.94) |

PCP=Primary Care Provider, PA=Physician Assistant, DO=Doctor of Osteopathy, DC=Doctor of Chiropractic, PT=Physical Therapist, LAc=Licensed Acupuncturist, OS=Orthpedic Surgeon, PMR=Physical Medicine & Rehabilitation, PM=Pain Management, NS=Neurosurgeon, Neuro=Neurologist, Rheum=Rheumatologist, MD Oth=Other MD specialty, EM=Emergency Medicine, Rad=Radiologist, UC=Urgent [illegible]

Cells in red are not different than the reference sement of 50-75 ADI and 75-100% NHW (p=.05)
