## Supplement 5 - Services for "Geographic variation in the treatment of spinal disorders: association with health care professional availability, and population socioeconomic status, race, and ethnicity. A retrospective cohort study"

| Supplement 5 - % of spinal disorder episodes including health care service | | | | | | | | | | | | | | | | | | | |
|---|---|---|---|---|---|---|---|---|---|---|---|---|---|---|---|---|---|---|---|
| ADI | % NHW | Episodes | Any | | | First Line | | | | | | Second Line | | | | Third Line | | | |
| | | | First Line | Second Line | Third Line | CMT | AC | PT | MT | Acu | OMT | Rad | NSAID | MMRelax | MRI | Opioid | Inj | Surg | CT |
| Total | | 1510869 | 45.0% | 52.7% | 21.5% | 33.6% | 19.8% | 15.9% | 14.1% | 1.2% | 0.9% | 25.3% | 22.4% | 21.1% | 10.1% | 15.5% | 5.9% | 3.3% | 3.2% |
| 75-100 | All | 128401 | 32.7% | 60.0% | 25.4% | 24.8% | 12.2% | 11.9% | 8.4% | 0.2% | 0.7% | 27.7% | 28.9% | 26.2% | 9.1% | 19.0% | 5.4% | 3.2% | 4.5% |
| 50-75 | | 500526 | 41.9% | 54.7% | 23.3% | 32.9% | 15.3% | 14.6% | 10.7% | 0.5% | 0.8% | 26.0% | 24.1% | 22.9% | 9.4% | 17.3% | 5.7% | 3.4% | 3.6% |
| 25-50 | | 557917 | 47.4% | 51.3% | 21.0% | 35.8% | 20.7% | 16.4% | 14.3% | 1.1% | 0.8% | 24.9% | 21.1% | 20.5% | 10.0% | 15.1% | 6.0% | 3.3% | 2.9% |
| 0-25 | | 324025 | 50.6% | 48.9% | 18.1% | 34.2% | 28.3% | 18.9% | 21.4% | 3.1% | 1.0% | 24.0% | 19.4% | 17.2% | 11.5% | 12.2% | 6.1% | 3.1% | 2.4% |
| All | 0-25 | 125079 | 28.4% | 63.6% | 21.4% | 17.3% | 16.6% | 12.0% | 11.9% | 1.8% | 0.5% | 28.1% | 33.8% | 26.4% | 9.4% | 15.0% | 4.8% | 2.2% | 4.5% |
| | 25-50 | 223789 | 38.2% | 57.7% | 22.1% | 25.6% | 20.5% | 15.2% | 14.7% | 1.7% | 0.7% | 26.8% | 26.8% | 23.5% | 10.2% | 16.0% | 5.7% | 2.9% | 3.5% |
| | 50-75 | 461777 | 43.8% | 54.0% | 22.4% | 31.2% | 21.9% | 16.5% | 15.7% | 1.5% | 0.9% | 26.0% | 22.8% | 21.6% | 10.7% | 16.2% | 6.3% | 3.4% | 3.1% |
| | 75-100 | 700224 | 50.9% | 48.2% | 20.8% | 40.6% | 18.7% | 16.5% | 13.3% | 0.8% | 0.9% | 23.9% | 18.7% | 19.0% | 9.7% | 15.1% | 5.8% | 3.5% | 2.9% |
| 75-100 | 0-25 | 35387 | 23.4% | 66.6% | 24.0% | 14.2% | 13.3% | 9.4% | 8.4% | 0.3% | 0.6% | 29.5% | 35.4% | 29.4% | 8.5% | 17.2% | 4.7% | 2.3% | 5.1% |
| | 25-50 | 24343 | 29.8% | 62.3% | 26.6% | 21.2% | 13.2% | 11.4% | 8.8% | 0.3% | 0.7% | 29.0% | 30.3% | 27.1% | 9.5% | 19.7% | 5.8% | 3.1% | 4.7% |
| | 50-75 | 27185 | 31.8% | 61.0% | 27.8% | 24.4% | 11.9% | 12.2% | 8.5% | 0.2% | 0.7% | 28.4% | 28.4% | 27.0% | 9.7% | 21.6% | 5.9% | 3.7% | 4.4% |
| | 75-100 | 41486 | 43.0% | 52.5% | 24.2% | 36.4% | 10.9% | 14.2% | 8.0% | 0.1% | 0.9% | 25.1% | 22.8% | 22.5% | 9.0% | 18.4% | 5.2% | 3.6% | 3.9% |
| 50-75 | 0-25 | 49075 | 27.0% | 65.1% | 22.3% | 16.9% | 15.6% | 11.2% | 10.2% | 0.9% | 0.5% | 28.7% | 34.8% | 27.6% | 9.1% | 16.0% | 4.8% | 2.3% | 4.5% |
| | 25-50 | 84091 | 34.7% | 60.1% | 24.4% | 24.0% | 16.9% | 13.5% | 11.7% | 0.7% | 0.7% | 27.7% | 28.1% | 25.6% | 10.1% | 18.0% | 6.1% | 3.1% | 3.9% |
| | 50-75 | 139453 | 38.8% | 57.2% | 25.1% | 29.3% | 16.3% | 14.5% | 11.1% | 0.4% | 0.8% | 27.4% | 25.0% | 24.3% | 10.1% | 18.7% | 6.4% | 3.7% | 3.6% |
| | 75-100 | 227907 | 49.6% | 48.8% | 22.0% | 41.8% | 14.0% | 15.7% | 10.1% | 0.3% | 0.9% | 24.0% | 19.8% | 20.1% | 8.8% | 16.5% | 5.4% | 3.5% | 3.3% |
| 25-50 | 0-25 | 29006 | 31.2% | 61.6% | 18.7% | 19.6% | 18.5% | 13.1% | 14.2% | 2.3% | 0.5% | 27.1% | 32.7% | 23.7% | 10.4% | 12.4% | 4.9% | 2.2% | 4.3% |
| | 25-50 | 68794 | 40.2% | 57.0% | 21.4% | 26.9% | 22.5% | 15.8% | 15.3% | 1.5% | 0.7% | 26.5% | 26.4% | 22.9% | 10.2% | 15.4% | 5.7% | 2.8% | 3.2% |
| | 50-75 | 176075 | 44.5% | 53.8% | 22.5% | 31.8% | 22.4% | 16.6% | 15.4% | 1.2% | 0.9% | 25.9% | 22.3% | 21.9% | 10.7% | 16.2% | 6.5% | 3.5% | 3.1% |
| | **75-100 - ref** | **284042** | **52.5%** | **47.4%** | **20.2%** | **42.1%** | **19.4%** | **16.8%** | **13.5%** | **0.8%** | **0.9%** | **23.6%** | **17.8%** | **18.8%** | **9.5%** | **14.7%** | **5.8%** | **3.5%** | **2.6%** |
| 0-25 | 0-25 | 11611 | 42.0% | 53.0% | 16.0% | 22.6% | 26.2% | 20.2% | 23.5% | 8.9% | 0.3% | 23.4% | 26.9% | 19.2% | 10.2% | 11.0% | 4.2% | 2.0% | 2.6% |
| | 25-50 | 46561 | 45.9% | 51.8% | 16.6% | 28.9% | 27.8% | 19.1% | 22.5% | 4.7% | 0.7% | 24.4% | 23.1% | 18.5% | 10.8% | 11.3% | 5.1% | 2.3% | 2.4% |
| | 50-75 | 119064 | 51.3% | 48.9% | 17.9% | 33.9% | 30.2% | 19.8% | 23.2% | 3.5% | 1.1% | 24.1% | 19.6% | 16.9% | 11.5% | 12.0% | 6.1% | 3.0% | 2.3% |
| | 75-100 | 146789 | 52.2% | 47.8% | 19.0% | 37.1% | 27.0% | 17.9% | 19.3% | 1.9% | 1.1% | 23.9% | 17.5% | 17.0% | 11.8% | 12.7% | 6.6% | 3.6% | 2.4% |

ADI=Area Deprivation Index, NHW=Non-Hispanic White, CMT=Chiropractic Manipulative Therapy, AC=Active Care, PT=Passive Therapy, MT=Manual Therapy, Acu=Acupuncture, OMT=Osteopathic Manipulative Therapy, Rad=Radiology, NSAID=prescription Non-Steroidal Anti-Inflammatory medication, MMRelax=prescription skeletal muslce relaxant medication, MRI=Magnetic Resonance Imaging, Opioid=prescription opioid medication, Inj=spinal injection, Surg=spinal surgery, CT=Computed Tomography scan

Cells with red text denote that the effect of provider type on service usage was found not to be significantly different from that of ADI 0-25 % NHW 75-100 reference (Fisher's Exact $p > 0.001$)
Cells with black text denote that the effect of provider type on service usage was found to be significantly different from that of ADI 0-25 % NHW 75-100 reference (Fisher's Exact $p < 0.001$)
