## Supplement 5a - Services Risk Ratio for "Geographic variation in the treatment of spinal disorders: association with health care professional availability, and population socioeconomic status, race, and ethnicity. A retrospective cohort study"

### Supplement 5a - Risk ratio and 95% confidence interval for % of episodes including service compared to reference segment of 25-50 Area Deprivation Index (ADI) and 75-100% non-Hispanic white (NHW) population

| ADI | % NHW | Any | | | First Line | | | | | | Second Line | | | | Third Line | | | |
|---|---|---|---|---|---|---|---|---|---|---|---|---|---|---|---|---|---|---|
| | | First Line | Second Line | Third Line | CMT | AC | PT | MT | Acu | OMT | Rad | NSAID | MM Relax | MRI | Opioid | Inj | Surg | CT |
| 75-100 | 0-25 | 0.45 (0.44, 0.45) | 1.41 (1.40, 1.42) | 1.19 (1.16, 1.21) | 0.34 (0.33, 0.35) | 0.68 (0.66, 0.70) | 0.56 (0.54, 0.58) | 0.62 (0.60, 0.64) | 0.41 (0.34, 0.49) | 0.68 (0.59, 0.78) | 1.25 (1.23, 1.27) | 1.99 (1.95, 2.02) | 1.57 (1.54, 1.59) | 0.90 (0.86, 0.93) | 1.18 (1.15, 1.20) | 0.81 (0.77, 0.85) | 0.66 (0.61, 0.70) | 1.94 (1.84, 2.04) |
| | 25-50 | 0.57 (0.56, 0.58) | 1.32 (1.30, 1.33) | 1.31 (1.28, 1.34) | 0.50 (0.49, 0.52) | 0.68 (0.66, 0.70) | 0.68 (0.66, 0.71) | 0.66 (0.63, 0.68) | 0.36 (0.28, 0.46) | 0.76 (0.65, 0.90) | 1.23 (1.20, 1.25) | 1.70 (1.67, 1.74) | 1.45 (1.41, 1.48) | 1.00 (0.96, 1.04) | 1.34 (1.31, 1.38) | 1.00 (0.95, 1.05) | 0.90 (0.84, 0.97) | 1.79 (1.69, 1.90) |
| | 50-75 | 0.61 (0.60, 0.62) | 1.29 (1.27, 1.30) | 1.38 (1.35, 1.40) | 0.58 (0.57, 0.59) | 0.61 (0.59, 0.63) | 0.73 (0.70, 0.75) | 0.63 (0.61, 0.66) | 0.23 (0.18, 0.31) | 0.84 (0.73, 0.97) | 1.20 (1.18, 1.23) | 1.59 (1.56, 1.63) | 1.44 (1.41, 1.47) | 1.02 (0.98, 1.06) | 1.47 (1.44, 1.51) | 1.01 (0.96, 1.06) | 1.06 (0.99, 1.13) | 1.67 (1.57, 1.77) |
| | 75-100 | 0.82 (0.81, 0.83) | 1.11 (1.10, 1.12) | 1.20 (1.18, 1.22) | 0.86 (0.85, 0.88) | 0.56 (0.55, 0.58) | 0.85 (0.83, 0.87) | 0.60 (0.58, 0.62) | 0.17 (0.13, 0.23) | 1.11 (1.00, 1.23) | 1.06 (1.05, 1.08) | 1.28 (1.26, 1.31) | 1.20 (1.18, 1.22) | 0.95 (0.92, 0.98) | 1.25 (1.22, 1.28) | 0.90 (0.86, 0.94) | 1.03 (0.98, 1.09) | 1.48 (1.40, 1.56) |
| 50-75 | 0-25 | 0.51 (0.51, 0.52) | 1.37 (1.36, 1.39) | 1.10 (1.08, 1.12) | 0.40 (0.39, 0.41) | 0.80 (0.78, 0.82) | 0.67 (0.65, 0.69) | 0.76 (0.74, 0.78) | 1.15 (1.04, 1.28) | 0.60 (0.53, 0.68) | 1.22 (1.20, 1.24) | 1.96 (1.93, 1.99) | 1.47 (1.45, 1.49) | 0.96 (0.93, 0.99) | 1.09 (1.06, 1.11) | 0.83 (0.79, 0.86) | 0.66 (0.62, 0.70) | 1.73 (1.65, 1.81) |
| | 25-50 | 0.66 (0.65, 0.67) | 1.27 (1.26, 1.28) | 1.21 (1.19, 1.22) | 0.57 (0.56, 0.58) | 0.87 (0.85, 0.88) | 0.80 (0.79, 0.82) | 0.87 (0.85, 0.89) | 0.83 (0.75, 0.91) | 0.84 (0.77, 0.92) | 1.18 (1.16, 1.19) | 1.58 (1.56, 1.60) | 1.36 (1.35, 1.38) | 1.06 (1.03, 1.08) | 1.23 (1.21, 1.25) | 1.04 (1.01, 1.07) | 0.89 (0.85, 0.93) | 1.48 (1.42, 1.54) |
| | 50-75 | 0.74 (0.73, 0.74) | 1.21 (1.20, 1.22) | 1.24 (1.22, 1.25) | 0.70 (0.69, 0.70) | 0.84 (0.83, 0.85) | 0.87 (0.85, 0.88) | 0.83 (0.81, 0.84) | 0.53 (0.49, 0.59) | 0.89 (0.83, 0.95) | 1.16 (1.15, 1.18) | 1.40 (1.39, 1.42) | 1.29 (1.28, 1.31) | 1.07 (1.05, 1.09) | 1.27 (1.26, 1.29) | 1.09 (1.06, 1.12) | 1.06 (1.02, 1.09) | 1.38 (1.33, 1.43) |
| | 75-100 | 0.95 (0.94, 0.95) | 1.03 (1.03, 1.04) | 1.09 (1.08, 1.10) | 0.99 (0.99, 1.00) | 0.72 (0.71, 0.73) | 0.94 (0.92, 0.95) | 0.75 (0.74, 0.76) | 0.42 (0.39, 0.46) | 1.03 (0.98, 1.10) | 1.02 (1.01, 1.03) | 1.11 (1.10, 1.13) | 1.07 (1.06, 1.08) | 0.92 (0.91, 0.94) | 1.13 (1.11, 1.14) | 0.92 (0.90, 0.94) | 1.00 (0.98, 1.03) | 1.24 (1.20, 1.28) |
| 25-50 | 0-25 | 0.59 (0.58, 0.60) | 1.30 (1.29, 1.31) | 0.92 (0.90, 0.95) | 0.47 (0.46, 0.48) | 0.95 (0.93, 0.97) | 0.78 (0.76, 0.81) | 1.06 (1.03, 1.09) | 2.97 (2.73, 3.23) | 0.63 (0.54, 0.74) | 1.15 (1.13, 1.17) | 1.84 (1.81, 1.87) | 1.26 (1.23, 1.29) | 1.10 (1.06, 1.14) | 0.84 (0.82, 0.87) | 0.83 (0.79, 0.87) | 0.64 (0.59, 0.69) | 1.63 (1.54, 1.73) |
| | 25-50 | 0.77 (0.76, 0.77) | 1.20 (1.20, 1.21) | 1.06 (1.04, 1.07) | 0.64 (0.63, 0.65) | 1.16 (1.14, 1.17) | 0.94 (0.93, 0.96) | 1.14 (1.12, 1.16) | 1.93 (1.79, 2.07) | 0.78 (0.71, 0.86) | 1.13 (1.11, 1.14) | 1.49 (1.46, 1.51) | 1.22 (1.20, 1.24) | 1.07 (1.04, 1.09) | 1.05 (1.03, 1.07) | 0.98 (0.95, 1.01) | 0.81 (0.77, 0.85) | 1.21 (1.16, 1.27) |
| | 50-75 | 0.85 (0.84, 0.85) | 1.14 (1.13, 1.14) | 1.11 (1.10, 1.12) | 0.76 (0.75, 0.76) | 1.15 (1.14, 1.16) | 0.99 (0.97, 1.00) | 1.14 (1.13, 1.16) | 1.47 (1.38, 1.56) | 1.10 (1.03, 1.17) | 1.10 (1.09, 1.11) | 1.25 (1.24, 1.27) | 1.17 (1.15, 1.18) | 1.13 (1.11, 1.15) | 1.10 (1.09, 1.12) | 1.12 (1.09, 1.14) | 1.00 (0.97, 1.04) | 1.16 (1.12, 1.20) |
| | 75-100 | reference | | | | | | | | | | | | | | | | |
| 0-25 | 0-25 | 0.80 (0.78, 0.82) | 1.12 (1.10, 1.14) | 0.79 (0.76, 0.82) | 0.54 (0.52, 0.56) | 1.35 (1.31, 1.39) | 1.20 (1.16, 1.25) | 1.75 (1.69, 1.81) | 11.28 (10.50, 12.11) | 0.33 (0.24, 0.47) | 0.99 (0.96, 1.03) | 1.51 (1.47, 1.56) | 1.02 (0.98, 1.06) | 1.08 (1.02, 1.14) | 0.75 (0.71, 0.79) | 0.72 (0.66, 0.78) | 0.57 (0.51, 0.65) | 0.98 (0.87, 1.10) |
| | 25-50 | 0.87 (0.86, 0.88) | 1.09 (1.08, 1.10) | 0.82 (0.80, 0.84) | 0.69 (0.68, 0.70) | 1.43 (1.40, 1.45) | 1.14 (1.12, 1.16) | 1.67 (1.64, 1.70) | 6.04 (5.70, 6.40) | 0.81 (0.72, 0.90) | 1.03 (1.02, 1.05) | 1.30 (1.27, 1.32) | 0.99 (0.97, 1.01) | 1.13 (1.10, 1.17) | 0.77 (0.75, 0.79) | 0.86 (0.83, 0.90) | 0.67 (0.63, 0.71) | 0.91 (0.85, 0.97) |
| | 50-75 | 0.98 (0.97, 0.98) | 1.03 (1.02, 1.04) | 0.88 (0.87, 0.90) | 0.80 (0.80, 0.81) | 1.55 (1.54, 1.57) | 1.18 (1.16, 1.19) | 1.72 (1.70, 1.75) | 4.49 (4.27, 4.73) | 1.25 (1.16, 1.33) | 1.02 (1.01, 1.03) | 1.10 (1.08, 1.11) | 0.90 (0.88, 0.91) | 1.21 (1.19, 1.24) | 0.82 (0.80, 0.83) | 1.05 (1.02, 1.07) | 0.87 (0.84, 0.91) | 0.86 (0.83, 0.90) |
| | 75-100 | 0.99 (0.99, 1.00) | 1.01 (1.00, 1.02) | 0.94 (0.93, 0.95) | 0.88 (0.87, 0.89) | 1.39 (1.37, 1.41) | 1.07 (1.05, 1.08) | 1.44 (1.42, 1.46) | 2.40 (2.27, 2.53) | 1.34 (1.26, 1.42) | 1.01 (1.00, 1.02) | 0.98 (0.97, 1.00) | 0.90 (0.89, 0.92) | 1.24 (1.22, 1.26) | 0.86 (0.85, 0.88) | 1.13 (1.10, 1.15) | 1.02 (0.99, 1.06) | 0.91 (0.87, 0.94) |

ADI=Area Deprivation Index, NHW=Non-Hispanic White, CMT=Chiropractic Manipulative Therapy, AC=Active Care, PT=Passive Therapy, MT=Manual Therapy, Acu=Acupuncture, OMT=Osteopathic Manipulative Therapy, Rad=Radiology, NSAID=prescription Non-Steroidal Anti-Inflammatory medication, MMRelax=prescription skeletal muslce relaxant medication, MRI=Magnetic Resonance Imaging, Opioid=prescription opioid medication, Inj=spinal injection, Surg=spinal surgery, CT=Computed Tomography scan

Cells in red are not different than the reference sement of 50-75 ADI and 75-100% NHW (p=.05)
