## Supplement 6 - Services Timing for "Geographic variation in the treatment of spinal disorders: association with health care professional availability, and population socioeconomic status, race, and ethnicity. A retrospective cohort study"

**Supplement 6 - # of days into episode when service first provided - Median (Q1, Q3)**

| ADI | % NHW | Episodes | Any | | | First Line | | | | | | Second Line | | | | Third Line | | | |
|---|---|---|---|---|---|---|---|---|---|---|---|---|---|---|---|---|---|---|---|
| | | | First Line | Second Line | Third Line | CMT | AC | PT | MT | Acu | OMT | Rad | NSAID | MMRelax | MRI | Opioid | Inj | Surg | CT |
| Total | | 1510869 | 0 (0, 20) | 0 (0, 39) | 6 (0, 72) | 0 (0, 0) | 4 (0, 32) | 0 (0, 21) | 6 (0, 37) | 0 (0, 19) | 0 (0, 19) | 0 (0, 19) | 0 (0, 29) | 0 (0, 14) | 20 (3, 78) | 1 (0, 50) | 39 (7, 107) | 44 (14, 106) | 0 (0, 40) |
| 75-100 | All | 128401 | 0 (0, 21) | 0 (0, 39) | 2 (0, 65) | 0 (0, 0) | 8 (0, 45) | 0 (0, 27) | 8 (0, 44) | 0 (0, 39) | 0 (0, 10) | 0 (0, 19) | 0 (0, 22) | 0 (0, 15) | 24 (4, 88) | 1 (0, 45) | 43 (8, 114) | 49 (15, 111) | 0 (0, 27) |
| 50-75 | All | 500526 | 0 (0, 19) | 0 (0, 40) | 4 (0, 71) | 0 (0, 0) | 7 (0, 41) | 0 (0, 22) | 7 (0, 44) | 0 (0, 35) | 0 (0, 17) | 0 (0, 20) | 0 (0, 28) | 0 (0, 15) | 22 (4, 84) | 1 (0, 48) | 43 (8, 114) | 49 (15, 112) | 0 (0, 38) |
| 25-50 | All | 557917 | 0 (0, 19) | 0 (0, 39) | 7 (0, 74) | 0 (0, 0) | 4 (0, 31) | 0 (0, 21) | 6 (0, 38) | 0 (0, 27) | 0 (0, 21) | 0 (0, 18) | 0 (0, 30) | 0 (0, 13) | 20 (3, 78) | 1 (0, 52) | 38 (7, 105) | 43 (13, 103) | 0 (0, 42) |
| 0-25 | All | 324025 | 0 (0, 22) | 0 (0, 37) | 10 (0, 77) | 0 (0, 0) | 2 (0, 24) | 0 (0, 18) | 3 (0, 29) | 0 (0, 10) | 0 (0, 19) | 0 (0, 16) | 0 (0, 31) | 0 (0, 10) | 17 (3, 69) | 1 (0, 56) | 35 (6, 99) | 39 (11, 98) | 0 (0, 50) |
| All | 0-25 | 125079 | 1 (0, 28) | 0 (0, 28) | 1 (0, 61) | 0 (0, 2) | 5 (0, 33) | 0 (0, 21) | 6 (0, 36) | 0 (0, 4) | 0 (0, 18) | 0 (0, 11) | 0 (0, 10) | 0 (0, 7) | 20 (3, 74) | 1 (0, 44) | 38 (5, 106) | 48 (14, 109) | 0 (0, 17) |
| All | 25-50 | 223789 | 0 (0, 25) | 0 (0, 33) | 4 (0, 68) | 0 (0, 1) | 3 (0, 30) | 0 (0, 21) | 5 (0, 35) | 0 (0, 13) | 0 (0, 20) | 0 (0, 14) | 0 (0, 21) | 0 (0, 9) | 20 (3, 76) | 1 (0, 47) | 37 (7, 107) | 46 (14, 111) | 0 (0, 31) |
| All | 50-75 | 461777 | 0 (0, 22) | 0 (0, 39) | 7 (0, 74) | 0 (0, 0) | 3 (0, 30) | 0 (0, 21) | 5 (0, 35) | 0 (0, 16) | 0 (0, 21) | 0 (0, 18) | 0 (0, 30) | 0 (0, 13) | 20 (3, 76) | 1 (0, 51) | 38 (7, 105) | 44 (13, 105) | 0 (0, 46) |
| All | 75-100 | 700224 | 0 (0, 15) | 0 (0, 44) | 7 (0, 76) | 0 (0, 0) | 5 (0, 35) | 0 (0, 21) | 7 (0, 40) | 0 (0, 33) | 0 (0, 17) | 0 (0, 22) | 0 (0, 38) | 0 (0, 17) | 21 (4, 81) | 1 (0, 52) | 41 (8, 110) | 43 (14, 105) | 0 (0, 48) |
| 75-100 | 0-25 | 35387 | 2 (0, 31) | 0 (0, 26) | 1 (0, 56) | 0 (0, 4) | 7 (0, 40) | 0 (0, 25) | 9 (0, 43) | 0 (0, 26) | 0 (0, 9) | 0 (0, 11) | 0 (0, 9) | 0 (0, 6) | 22 (3, 83) | 1 (0, 41) | 41 (5, 108) | 48 (12, 107) | 0 (0, 12) |
| 75-100 | 25-50 | 24343 | 0 (0, 28) | 0 (0, 35) | 2 (0, 63) | 0 (0, 1) | 8 (0, 44) | 0 (0, 29) | 10 (0, 42) | 0 (0, 27) | 0 (0, 10) | 0 (0, 16) | 0 (0, 19) | 0 (0, 12) | 24 (4, 86) | 1 (0, 47) | 41 (9, 111) | 45 (15, 108) | 0 (0, 20) |
| 75-100 | 50-75 | 27185 | 0 (0, 23) | 0 (0, 45) | 2 (0, 68) | 0 (0, 1) | 8 (0, 47) | 0 (0, 28) | 7 (0, 43) | 5 (0, 73) | 0 (0, 12) | 0 (0, 24) | 0 (0, 30) | 0 (0, 20) | 25 (5, 89) | 1 (0, 42) | 43 (8, 122) | 46 (14, 111) | 0 (0, 37) |
| 75-100 | 75-100 | 41486 | 0 (0, 12) | 0 (0, 52) | 4 (0, 74) | 0 (0, 0) | 8 (0, 51) | 0 (0, 25) | 7 (0, 47) | 9 (0, 53) | 0 (0, 8) | 0 (0, 28) | 0 (0, 38) | 0 (0, 26) | 26 (6, 93) | 1 (0, 49) | 46 (8, 117) | 52 (16, 113) | 0 (0, 45) |
| 50-75 | 0-25 | 49075 | 1 (0, 28) | 0 (0, 27) | 1 (0, 60) | 0 (0, 3) | 6 (0, 35) | 0 (0, 21) | 8 (0, 42) | 0 (0, 14) | 0 (0, 18) | 0 (0, 11) | 0 (0, 11) | 0 (0, 7) | 20 (3, 74) | 1 (0, 42) | 42 (8, 108) | 51 (17, 111) | 0 (0, 13) |
| 50-75 | 25-50 | 84091 | 0 (0, 27) | 0 (0, 35) | 4 (0, 66) | 0 (0, 1) | 6 (0, 37) | 0 (0, 23) | 7 (0, 42) | 0 (0, 32) | 0 (0, 19) | 0 (0, 15) | 0 (0, 22) | 0 (0, 11) | 21 (3, 80) | 1 (0, 45) | 38 (7, 109) | 46 (14, 109) | 0 (0, 36) |
| 50-75 | 50-75 | 139453 | 0 (0, 23) | 0 (0, 40) | 5 (0, 72) | 0 (0, 1) | 7 (0, 40) | 0 (0, 23) | 8 (0, 44) | 0 (0, 33) | 0 (0, 19) | 0 (0, 20) | 0 (0, 29) | 0 (0, 15) | 22 (4, 83) | 1 (0, 48) | 41 (8, 112) | 48 (15, 112) | 0 (0, 41) |
| 50-75 | 75-100 | 227907 | 0 (0, 12) | 0 (0, 47) | 6 (0, 74) | 0 (0, 0) | 7 (0, 46) | 0 (0, 22) | 7 (0, 47) | 4 (0, 61) | 0 (0, 15) | 0 (0, 26) | 0 (0, 39) | 0 (0, 20) | 24 (5, 88) | 1 (0, 50) | 46 (10, 119) | 49 (15, 113) | 0 (0, 46) |
| 25-50 | 0-25 | 29006 | 1 (0, 27) | 0 (0, 28) | 3 (0, 65) | 0 (0, 2) | 2 (0, 27) | 0 (0, 20) | 4 (0, 31) | 0 (0, 12) | 0 (0, 21) | 0 (0, 12) | 0 (0, 11) | 0 (0, 7) | 18 (2, 68) | 1 (0, 51) | 30 (1, 98) | 42 (10, 106) | 0 (0, 29) |
| 25-50 | 25-50 | 68794 | 0 (0, 24) | 0 (0, 32) | 5 (0, 69) | 0 (0, 1) | 2 (0, 28) | 0 (0, 20) | 5 (0, 35) | 0 (0, 18) | 0 (0, 21) | 0 (0, 14) | 0 (0, 21) | 0 (0, 8) | 19 (3, 72) | 1 (0, 48) | 38 (7, 105) | 49 (16, 113) | 0 (0, 30) |
| 25-50 | 50-75 | 176075 | 0 (0, 23) | 0 (0, 39) | 7 (0, 73) | 0 (0, 0) | 3 (0, 30) | 0 (0, 21) | 6 (0, 37) | 0 (0, 23) | 0 (0, 26) | 0 (0, 17) | 0 (0, 31) | 0 (0, 13) | 19 (3, 77) | 1 (0, 51) | 38 (7, 102) | 44 (13, 102) | 0 (0, 44) |
| 25-50 | **75-100 - ref** | **284042** | **0 (0, 15)** | **0 (0, 43)** | **8 (0, 77)** | **0 (0, 0)** | **5 (0, 34)** | **0 (0, 21)** | **7 (0, 41)** | **0 (0, 41)** | **0 (0, 16)** | **0 (0, 21)** | **0 (0, 38)** | **0 (0, 16)** | **21 (3, 82)** | **1 (0, 54)** | **40 (8, 107)** | **42 (13, 102)** | **0 (0, 48)** |
| 0-25 | 0-25 | 11611 | 0 (0, 28) | 0 (0, 31) | 5 (0, 76) | 0 (0, 0) | 2 (0, 27) | 0 (0, 17) | 0 (0, 27) | 0 (0, 0) | 0 (0, 7) | 0 (0, 10) | 0 (0, 17) | 0 (0, 4) | 18 (2, 69) | 1 (0, 49) | 42 (6, 120) | 55 (14, 110) | 0 (0, 39) |
| 0-25 | 25-50 | 46561 | 0 (0, 24) | 0 (0, 31) | 7 (0, 72) | 0 (0, 0) | 1 (0, 22) | 0 (0, 17) | 1 (0, 26) | 0 (0, 6) | 0 (0, 24) | 0 (0, 13) | 0 (0, 19) | 0 (0, 5) | 18 (3, 69) | 1 (0, 50) | 35 (6, 103) | 41 (11, 111) | 0 (0, 31) |
| 0-25 | 50-75 | 119064 | 0 (0, 22) | 0 (0, 37) | 11 (0, 78) | 0 (0, 0) | 1 (0, 22) | 0 (0, 17) | 2 (0, 28) | 0 (0, 10) | 0 (0, 15) | 0 (0, 16) | 0 (0, 32) | 0 (0, 10) | 17 (3, 67) | 2 (0, 58) | 34 (6, 98) | 39 (11, 97) | 0 (0, 58) |
| 0-25 | 75-100 | 146789 | 0 (0, 21) | 0 (0, 40) | 11 (0, 77) | 0 (0, 0) | 2 (0, 26) | 0 (0, 20) | 5 (0, 34) | 0 (0, 20) | 0 (0, 21) | 0 (0, 19) | 0 (0, 38) | 0 (0, 13) | 18 (3, 69) | 1 (0, 56) | 35 (7, 99) | 37 (11, 98) | 0 (0, 51) |

ADI=Area Deprivation Index, NHW=Non-Hispanic White, CMT=Chiropractic Manipulative Therapy, AC=Active Care, PT=Passive Therapy, MT=Manual Therapy, Acu=Acupuncture, OMT=Osteopathic Manipulative Therapy, Rad=Radiology, NSAID=prescription Non-Steroidal Anti-Inflammatory medication, MMRelax=prescription skeletal muslce relaxant medication, MRI=Magnetic Resonance Imaging, Opioid=prescription opioid medication, Inj=spinal injection, Surg=spinal surgery, CT=Computed Tomography scan

Cells with red text denote that the effect of provider type on service usage was found not to be significantly different from that of ADI 0-25 % NHW 75-100 reference (Mann-Whitney U p > 0.001)

Cells with black text denote that the effect of provider type on service usage was found to be significantly different from that of ADI 0-25 % NHW 75-100 reference (Mann-Whitney U p > 0.001)
