## Supplement 7 - Cost for "Geographic variation in the treatment of spinal disorders: association with health care professional availability, and population socioeconomic status, race, and ethnicity. A retrospective cohort study"

| Supplement 7 - Total episode of care attributes - Median (Q1, Q3) | | | | | |
|---|---|---|---|---|---|
| ADI | % NHW | Episodes | Total Episode Cost | Episode Duration | # of Health Care Providers Seen |
| Total | | 1510869 | 195 (65, 662) | 26 (1, 134) | 2 (1, 3) |
| 75-100 | All | 128401 | 160 (52, 604) | 22 (1, 142) | 2 (1, 3) |
| 50-75 | | 500526 | 175 (58, 602) | 25 (1, 140) | 2 (1, 3) |
| 25-50 | | 557917 | 198 (68, 648) | 27 (1, 132) | 2 (1, 3) |
| 0-25 | | 324025 | 244 (90, 807) | 28 (1, 126) | 2 (1, 3) |
| All | 0-25 | 125079 | 187 (56, 701) | 15 (1, 114) | 2 (1, 3) |
| | 25-50 | 223789 | 202 (65, 711) | 22 (1, 125) | 2 (1, 3) |
| | 50-75 | 461777 | 205 (69, 711) | 28 (1, 136) | 2 (1, 3) |
| | 75-100 | 700224 | 188 (65, 616) | 29 (1, 139) | 1 (1, 3) |
| 75-100 | 0-25 | 35387 | 168 (50, 678) | 13 (1, 114) | 2 (1, 3) |
| | 25-50 | 24343 | 171 (55, 648) | 22 (1, 139) | 2 (1, 3) |
| | 50-75 | 27185 | 165 (55, 626) | 29 (1, 157) | 2 (1, 3) |
| | 75-100 | 41486 | 144 (50, 515) | 29 (1, 156) | 2 (1, 3) |
| 50-75 | 0-25 | 49075 | 181 (54, 687) | 15 (1, 117) | 2 (1, 3) |
| | 25-50 | 84091 | 190 (60, 661) | 22 (1, 131) | 2 (1, 3) |
| | 50-75 | 139453 | 180 (61, 637) | 28 (1, 144) | 2 (1, 3) |
| | 75-100 | 227907 | 165 (55, 549) | 28 (1, 145) | 1 (1, 3) |
| 25-50 | 0-25 | 29006 | 197 (60, 695) | 17 (1, 112) | 2 (1, 3) |
| | 25-50 | 68794 | 203 (66, 710) | 22 (1, 121) | 2 (1, 3) |
| | 50-75 | 176075 | 200 (69, 677) | 27 (1, 134) | 2 (1, 3) |
| | **75-100 ref** | **284042** | **194 (68, 610)** | **29 (1, 135)** | **1 (1, 3)** |
| 0-25 | 0-25 | 11611 | 244 (83, 841) | 22 (1, 106) | 2 (1, 3) |
| | 25-50 | 46561 | 240 (85, 811) | 23 (1, 115) | 2 (1, 3) |
| | 50-75 | 119064 | 257 (94, 860) | 28 (1, 126) | 2 (1, 3) |
| | 75-100 | 146789 | 236 (90, 764) | 29 (1, 132) | 2 (1, 3) |

ADI=Area Deprivation Index, NHW=Non-Hispanic White

Cells with red text denote that the effect of provider type on service usage was found not to be significantly different from that of ADI 0-25 % NHW 75-100 reference (Mann-Whitney U p > 0.001)
Cells with black text denote that the effect of provider type on service usage was found to be significantly different from that of ADI 0-25 % NHW 75-100 reference (Mann-Whitney U p > 0.001)
