## Supplement 8 - Ridge Regression for "Geographic variation in the treatment of spinal disorders: association with health care professional availability, and population socioeconomic status, race, and ethnicity. A retrospective cohort study"

| Supplement 8 - Ridge regression coefficients (95% Confidence Intervals) | | | | | |
|---|---|---|---|---|---|
| Features | | Cost | Log Cost | Rx-Opioid | Rx-NSAID |
| (Intercept) | | 1152 (627, 1677) | 6.399 (6.183, 6.614) | 0.106 (0.077, 0.135) | 0.176 (0.144, 0.208) |
| # of health care providers (HCP) per 100 population | PCP | -48 (-263, 167) | -0.141 (-0.229, -0.052) | -0.001 (-0.013, 0.010) | 0.011 (-0.003, 0.024) |
| | Nurse | -31 (-182, 120) | -0.003 (-0.065, 0.059) | -0.002 (-0.011, 0.006) | 0.003 (-0.006, 0.012) |
| | PA | 410 (98, 722) | 0.045 (-0.083, 0.174) | 0.002 (-0.016, 0.019) | -0.021 (-0.041, -0.002) |
| | DO | -24 (-993, 945) | 0.069 (-0.329, 0.467) | 0.012 (-0.041, 0.066) | 0.014 (-0.045, 0.074) |
| | DC | -34 (-542, 475) | 0.214 (0.005, 0.423) | -0.032 (-0.060, -0.004) | -0.054 (-0.085, -0.022) |
| | PT | 257 (-172, 686) | 0.117 (-0.060, 0.293) | 0.012 (-0.012, 0.036) | -0.015 (-0.041, 0.011) |
| | LAc | -316 (-1271, 638) | -0.188 (-0.580, 0.204) | 0.010 (-0.042, 0.063) | -0.004 (-0.063, 0.054) |
| | OS | 340 (-595, 1276) | 0.691 (0.307, 1.075) | -0.047 (-0.099, 0.004) | -0.072 (-0.129, -0.015) |
| | PMR | -88 (-1097, 922) | 0.333 (-0.081, 0.748) | 0.017 (-0.039, 0.073) | 0.039 (-0.023, 0.100) |
| | PM | 443 (-244, 1130) | 0.543 (0.261, 0.825) | 0.050 (0.012, 0.088) | 0.041 (-0.001, 0.083) |
| | NS | -511 (-1339, 316) | -0.016 (-0.356, 0.324) | -0.011 (-0.057, 0.034) | -0.032 (-0.083, 0.018) |
| | Neuro | -141 (-989, 706) | -0.059 (-0.407, 0.290) | -0.004 (-0.051, 0.043) | -0.053 (-0.105, -0.001) |
| | Rheum | -447 (-1377, 482) | -0.189 (-0.570, 0.193) | -0.029 (-0.080, 0.023) | -0.020 (-0.077, 0.037) |
| | MD (Oth) | -92 (-261, 77) | 0.039 (-0.030, 0.109) | 0.004 (-0.006, 0.013) | 0.013 (0.003, 0.024) |
| | EM | 58 (-130, 247) | 0.028 (-0.049, 0.106) | -0.007 (-0.017, 0.004) | -0.007 (-0.019, 0.004) |
| | UC | -209 (-1010, 592) | 0.384 (0.054, 0.713) | 0.027 (-0.017, 0.071) | 0.004 (-0.045, 0.053) |
| | Rad | -118 (-582, 346) | -0.141 (-0.332, 0.050) | -0.012 (-0.037, 0.014) | -0.014 (-0.043, 0.014) |
| % of episodes with health care provider (HCP) as initial contact | PCP | -280 (-515, -45) | -0.074 (-0.170, 0.023) | 0.107 (0.094, 0.120) | 0.143 (0.129, 0.158) |
| | Nurse | -511 (-861, -160) | -0.138 (-0.282, 0.006) | 0.083 (0.064, 0.103) | 0.155 (0.134, 0.176) |
| | PA | 370 (-92, 832) | 0.103 (-0.087, 0.293) | 0.036 (0.011, 0.062) | 0.071 (0.043, 0.099) |
| | DO | -592 (-1504, 320) | 0.086 (-0.289, 0.461) | -0.086 (-0.137, -0.036) | -0.075 (-0.131, -0.019) |
| | DC | -927 (-1147, -708) | -0.349 (-0.439, -0.258) | -0.113 (-0.125, -0.100) | -0.167 (-0.180, -0.154) |
| | PT | -213 (-1170, 745) | 0.844 (0.450, 1.237) | -0.136 (-0.189, -0.083) | -0.038 (-0.097, 0.020) |
| | LAc | -430 (-1434, 574) | -0.196 (-0.608, 0.217) | -0.067 (-0.123, -0.012) | -0.163 (-0.224, -0.102) |
| | OS | 2949 (2437, 3461) | 1.654 (1.444, 1.864) | 0.072 (0.043, 0.100) | 0.151 (0.120, 0.182) |
| | PMR | 1624 (910, 2338) | 1.981 (1.688, 2.275) | 0.052 (0.013, 0.092) | -0.019 (-0.063, 0.025) |
| | PM | 483 (-447, 1414) | 1.753 (1.370, 2.135) | 0.259 (0.207, 0.310) | -0.013 (-0.070, 0.043) |
| | NS | 6029 (5146, 6912) | 3.633 (3.270, 3.996) | 0.184 (0.135, 0.233) | 0.025 (-0.029, 0.079) |
| | Neuro | 1711 (827, 2594) | 1.623 (1.261, 1.986) | 0.012 (-0.037, 0.061) | 0.020 (-0.034, 0.074) |
| | Rheum | 357 (-632, 1346) | 0.964 (0.557, 1.370) | 0.034 (-0.021, 0.089) | 0.084 (0.024, 0.145) |
| | MD (Oth) | 2645 (1870, 3420) | 0.562 (0.243, 0.880) | 0.039 (-0.004, 0.082) | 0.110 (0.063, 0.157) |
| | EM | 519 (-32, 1070) | 0.595 (0.368, 0.821) | 0.067 (0.036, 0.097) | 0.193 (0.160, 0.227) |
| | UC | -438 (-1457, 580) | 0.069 (-0.349, 0.488) | 0.014 (-0.042, 0.071) | -0.006 (-0.069, 0.056) |
| | Rad | 193 (-469, 854) | 0.590 (0.318, 0.862) | -0.135 (-0.172, -0.099) | -0.160 (-0.200, -0.119) |
| Zip code population attributes | ADI | -176 (-352, -0) | -0.647 (-0.719, -0.575) | 0.072 (0.062, 0.082) | 0.050 (0.039, 0.061) |
| | White | 328 (-131, 787) | 0.442 (0.253, 0.631) | 0.011 (-0.014, 0.036) | -0.022 (-0.050, 0.006) |
| | Black | -155 (-647, 336) | 0.688 (0.487, 0.890) | 0.005 (-0.022, 0.033) | 0.133 (0.103, 0.163) |
| | Asian | -70 (-759, 619) | 0.795 (0.512, 1.078) | -0.080 (-0.118, -0.042) | -0.006 (-0.048, 0.036) |
| | Hispanic | 503 (12, 993) | 0.927 (0.726, 1.129) | -0.014 (-0.041, 0.013) | 0.102 (0.072, 0.132) |

All features use similar measurement scale: HCP per 100, % initial provider, ADI 0-100, and % population race/ethnicity
Red cells represents p-value greater than threshold (0.05) - not significant
