## Supplement 8a - Ridge Regression Figure for "Geographic variation in the treatment of spinal disorders: association with health care professional availability, and population socioeconomic status, race, and ethnicity. A retrospective cohort study"

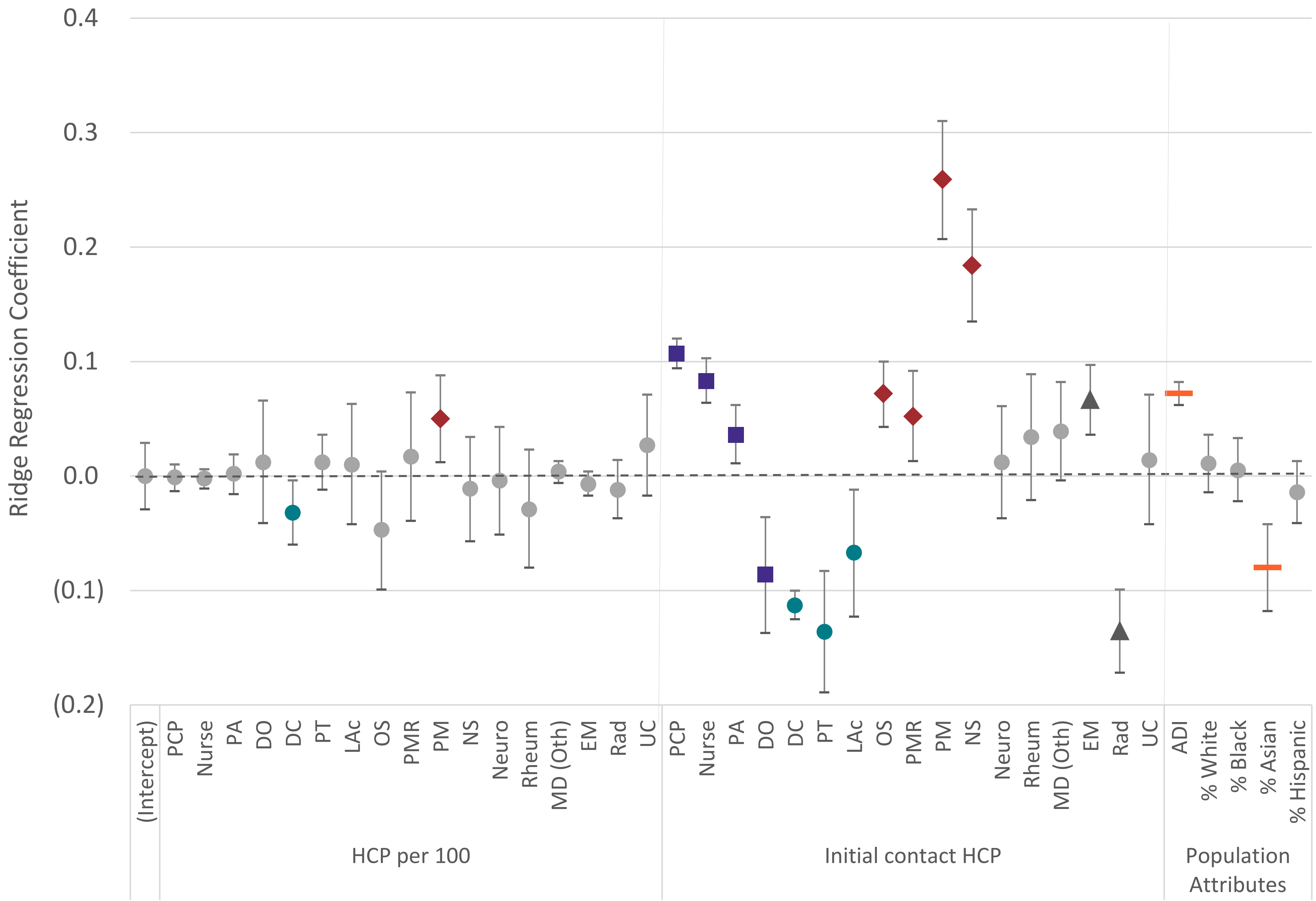


**Supplement 8a** - Ridge regression coefficients for association between health care provider (HCP) availability, initial contact HCP and population attributes on prescription opioid exposure for a spinal disorder episode
