## Supplement - Maps for "Geographic variation in the treatment of spinal disorders: association with health care professional availability, and population socioeconomic status, race, and ethnicity. A retrospective cohort study"

### # of Health Care Providers (HCP) Per 1000 Population - Chiropractors

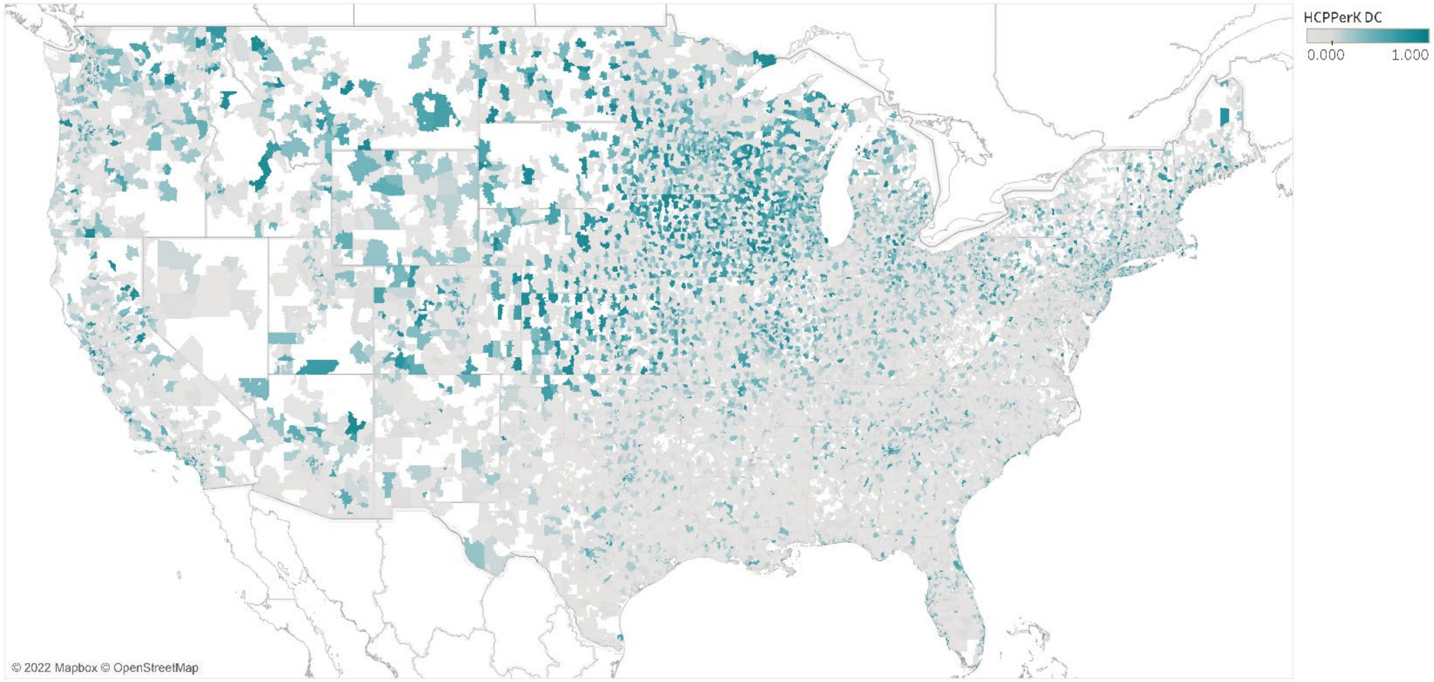


### % of Spinal Disorder Episodes Including Service Type - Rx-Opioid

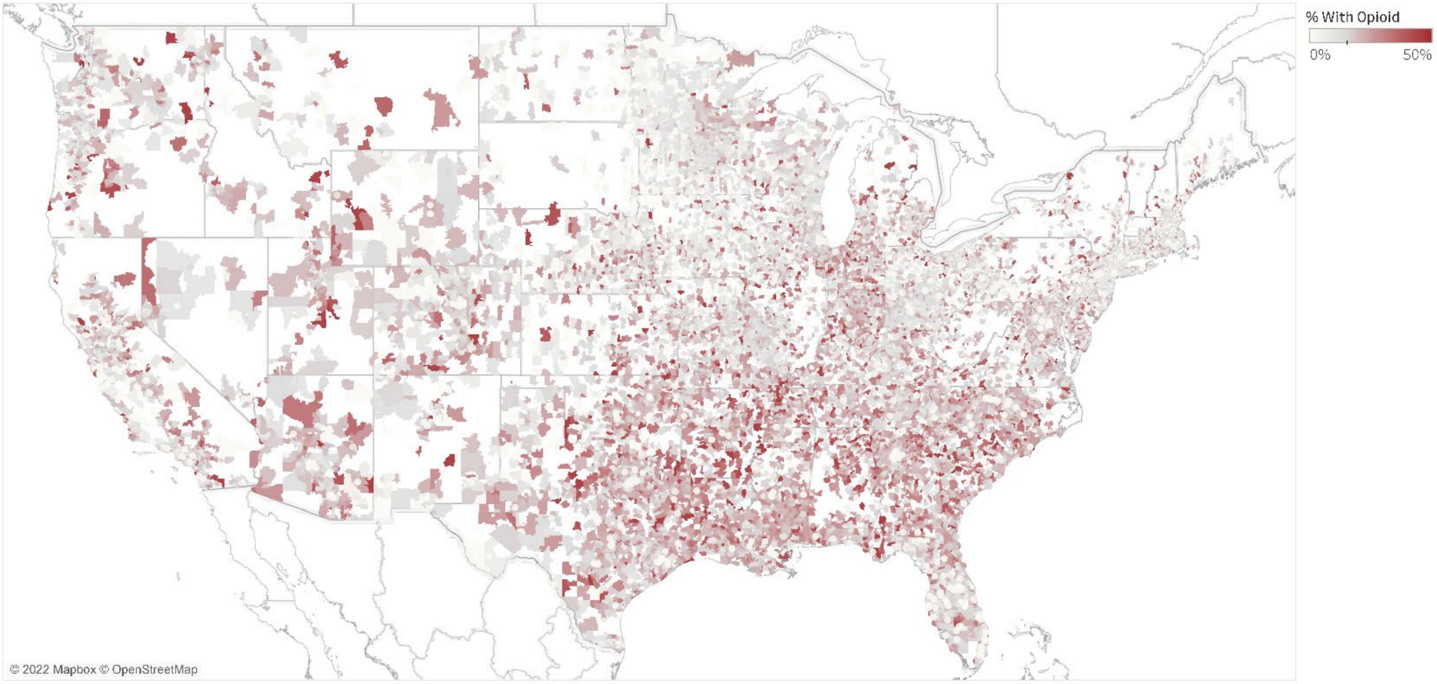
